# Data-driven modelling of tau pathology reveals distinct progressive supranuclear palsy subtypes

**DOI:** 10.1101/2025.08.03.25332866

**Authors:** Patrick W. Cullinane, Jacy Bezerra Parmera, Hemanth Nelvagal, Toby Curless, Leckhna Paras Chajed, Sarah Wrigley, Walter Sifontes Valladares, Maggie Burrows, Kirsten Ebanks, Lesley Wu, Lawrence P. Binding, Tamas Revesz, Raquel Real, David P. Vaughan, Edwin Jabbari, Huw R. Morris, Sebastian Brandner, Alexandra L. Young, Glen Hoti, Yangxinrui Ma, Mariam Bechtawi, Nancy Chiraki, Eduardo de Pablo-Fernández, Yau Mun Lim, Thomas T. Warner, Zane Jaunmuktane

**Affiliations:** Department of Clinical and Movement Neurosciences, UCL Queen Square Institute of Neurology, University College London, WC1N 3BG, UK; Queen Square Brain Bank for Neurological Disorders, UCL Queen Square Institute of Neurology, London, WC1N 1PJ, UK; Reta Lila Weston Institute of Neurological Studies, UCL Queen Square Institute of Neurology, London, WC1N 1PJ, UK; Queen Square Movement Disorders Centre, UCL Queen Square Institute of Neurology, London, WC1N 3BG, UK; Department of Neurology, Hospital das Clínicas, Faculdade de Medicina da Universidade de São Paulo (HC-FMUSP), São Paulo, 05402 000, Brazil; UCL Hawkes Institute and Department of Computer Science, University College London, London, WC1V 6LJ, UK; Department of Neurodegenerative Disease, UCL Queen Square Institute of Neurology, London, WC1N 3BG, UK; Division of Neuropathology, National Hospital for Neurology and Neurosurgery, University College London NHS Foundation Trust, London, WC1N 3BG, UK; Centre for Preventive Neurology, Wolfson Institute of Population Health, Queen Mary University London, EC1M 6BQ, UK

**Keywords:** Tauopathy, phenotypic heterogeneity, machine learning, disease progression modelling

## Abstract

Progressive supranuclear palsy (PSP) is a heterogeneous neurodegenerative disease characterised by the accumulation of misfolded 4-repeat tau within neurones and glial cells. There is limited longitudinal data on pathologically confirmed PSP patients with phenotypes other than classical Richardson’s syndrome (RS), and the pathomechanisms responsible for the broad variability in clinical phenotype and progression are not well understood. An unresolved question in this context is whether distinct spatiotemporal patterns of tau pathology propagation exist within the clinicopathological spectrum of PSP.

We identified 241 consecutive, pathologically confirmed patients with PSP from the Queen Square Brain Bank for Neurological Disorders (2010-2022). Phenotyping was performed based on clinical features present within the first 3 years from symptom onset according to the Movement Disorder Society (MDS) criteria, and specific clinical features and disease milestones were recorded. Genotyping was performed using Illumina NeuroBooster and NeuroChip arrays and *MAPT* haplotype, *APOE* genotype, *TRIM11* rs564309, and *SLC2A13* rs2242367 single nucleotide polymorphism status were collated from imputed data. Tissue sections from eight brain regions, mounted on glass slides, were immunostained for hyperphosphorylated tau and digitised using whole-slide scanning. Forty-one anatomical regions of interest were manually segmented, and total tau pathology burden was quantified using an automated, machine learning-based algorithm. The associations between survival and both clinicogenetic features and regional tau pathology burden were modelled using Cox regression and generalised linear models, respectively, and the Subtype and Stage Inference (SuStaIn) algorithm was used to identify subgroups with distinct progression patterns.

We have identified: 1) several clinical predictors of survival in PSP and the relationship between regional tau pathology burden and survival; 2) novel anatomical reference standards for the expected distribution of tau pathology across MDS-defined PSP phenotypes, emphasising region-specific white matter involvement in patients with corticobasal syndrome and speech/language variants; 3) associations linking biological sex, *MAPT* haplotype, and TDP-43 co-pathology to clinical phenotype and regional tau pathology burden; 4) patterns of covariance in regional tau pathology implicating inter-regional connectivity in tau spreading; and 5) three distinct spatiotemporal patterns of tau pathology progression: one characterised by initial involvement of subcortical grey matter followed by rostral spread to frontal white matter and other cortical regions, and two characterised by early, simultaneous involvement of subcortical grey matter and frontal white matter.

Taken together, these results indicate that PSP clinicopathological heterogeneity is mediated by propagation of tau pathology along anatomically connected networks, and via cell-autonomous mechanisms influenced by sex, genetic factors and possibly co-pathology.

## Introduction

Progressive supranuclear palsy (PSP) is characterised by the accumulation of misfolded 4- repeat tau within neurones and glial cells forming hallmark astrocytic inclusions in the latter, referred to as tufted astrocytes.^1,2^ It is a highly heterogeneous neurodegenerative disease with considerable variability in clinical phenotype and progression, leading to challenges for accurate early diagnosis, biomarker discovery and therapeutic trials.^3^ There is limited longitudinal data on pathologically confirmed PSP patients, particularly those with non- Richardson’s syndrome (RS) clinical phenotypes.^3,4^ While clinical phenotype broadly corresponds to the anatomical distribution of tau pathology,^5–10^ large-scale quantitative assessment of regional tau pathology across PSP clinical phenotypes is lacking, and the relationship between tau pathology burden in specific brain areas and clinical progression is not well understood. The relative contributions and interactions between cell autonomous (intrinsic susceptibility to protein misfolding) and non-autonomous (‘prion-like’ trans-synaptic spreading) mechanisms of tau pathology propagation in PSP also require further clarification. The current system for the pathological staging of PSP-RS emphasises a spatiotemporal pattern of tau propagation whereby tau accumulation begins in the pallido-nigro-luysian axis followed by subsequent spread rostrally to the neocortex and caudally to the cerebellum,^5^ while *in-vivo* work using [18F]Florzolotau PET imaging and data-driven modelling indicate that there is an additional pattern of tau pathology progression in some patients characterised by early simultaneous involvement of cortical and pallido-nigro-luysian regions.^11^ Limitations of previous clinicopathological studies include non-standardised clinical phenotyping,^12,13^ the use of semi-quantitative pathology assessments that are insensitive to subtle differences in tau pathology and do not account for tau burden in the hemispheric white matter,^5^ as well as comparatively small sample sizes.^14^ In this study, we aimed to evaluate clinical progression in a large cohort of clinically, genetically and pathologically well-characterised PSP patients, half of whom had clinical phenotypes other than PSP-RS based on clinical features present within the first 3 years from symptom onset. Using a machine learning-based digital pathology algorithm to generate quantitative measurements of regional tau pathology, we also aimed to determine patterns of tau pathology propagation through fine-grained clinicopathological correlation and data-driven disease progression modelling.^15^ The overall objective of this work is to uncover clinical, genetic and pathological factors that drive phenotypic diversity and disease progression in PSP in order to improve patient stratification, aid interpretation of biomarkers, and inform therapeutic interventions targeting removal of tau.

## Materials and methods

### Study design and participants

Patients with a neuropathological diagnosis of PSP were identified from the archives of the University College London (UCL) Queen Square Brain Bank for Neurological Disorders (QSBB) between 2010 and 2022. Consecutive patients were selected for inclusion, except where medical records did not contain adequate documentation of symptom onset, clinical features, and disease progression. QSBB protocols and this study have been approved by the NHS Health Research Authority Ethics Committee London-Central (REC reference 23/LO/0044). All donors included in this study or their next of kin gave written informed consent.

### Clinical assessment

All patients were regularly assessed by hospital specialists in the UK throughout the course of their illness. Patients’ primary and secondary care medical records were reviewed by neurologists with experience in movement disorders and using predefined definitions (supplementary data), we recorded demographic information, clinical features, and response to levodopa as previously described.^16^ The time from symptom onset to each clinical variable was calculated, and clinical predominance type (phenotype) was assigned based on the clinical features present within the first 3 years from symptom onset according to the Movement Disorder Society (MDS) PSP criteria^17^ and a modified version of the Multiple Allocations eXtinction (MAX) rules.^18^ The first MAX rule was omitted, thus favouring the predominance type rather than certainty level when assigning the phenotype to avoid overestimating PSP-RS, which is a criticism of this system.^19,20^ To evaluate disease progression, we recorded the time from symptom onset to specific disease milestones representing different domains of functional impairment. Milestones were chosen based on previous studies^13,21^and on the basis that they are likely to require medical attention and would be well documented in the medical records including: frequent falls and wheelchair dependence as markers of motor impairment; unintelligible speech/use of communication aids and the insertion or recommendation of percutaneous endoscopic gastrostomy (PEG) tube placement for feeding indicating severe bulbar dysfunction; dementia; and placement in residential/nursing home care or requirement of care for most activities of daily living in the home reflecting global disability. Records were available until patients’ death and the time from the onset of symptoms to death (survival) was also recorded. If a clinical feature/event was not recorded in the medical files, it was assumed absent. Ambiguity around the timing of clinical variables was resolved through consensus between raters.

### Genetics

DNA was extracted from cerebellar or frontal lobe tissue and genotyped using the Illumina NeuroBooster and NeuroChip arrays.^22,23^ Genotype data were filtered using PLINK v2^24^; individuals and SNPs with >5% missing data were excluded using –mind 0.05, –geno 0.05, – maf 0.01, and hwe1e-6 filters. Genotype data were imputed against the TOPMed r2 panel^25^ with Eagle v2.4^26^ phasing on the TOPMed Imputation Server using Minimac4. Variants previously reported to influence PSP tau pathology burden, clinical phenotype or progression were extracted from the imputed data, including *MAPT* haplotype (defined using rs1800547),^27,28^ the *TRIM11* SNP rs564309^29,30^ and the *SLC2A13* SNP rs2242367 located 190 Kb from the common *LRRK2* risk SNP for Parkinson’s disease.^31^ Patients homozygous (A/A) or heterozygous (C/A) for the *TRIM11* SNP rs564309 were designated *TRIM11* (A). Similarly, patients homozygous (A/A) or heterozygous (G/A) for the *SLC2A13* SNP rs2242367 were designated *SLC2A13* (A). Patients carrying one or two copies of the *MAPT* H2 haplotype (H1/H2 or H2/H2) were classified as *MAPT* H2. We also included *APOE* genotypes (defined using rs429358 and rs7412), given its association with Alzheimer’s disease (AD) pathology^32^ and PSP risk.^33^ Other genes associated with the risk of PSP were not included in this study.^34^

### Neuropathological examination

Pathological examination was performed following QSBB protocols. For histological analysis, 7 µm sections were cut from formalin-fixed, paraffin-embedded tissue blocks from representative brain regions. These included the anterior frontal lobe, encompassing the superior and middle frontal gyri; the posterior frontal lobe, containing the primary motor cortex; the temporal lobe, including the superior and middle temporal gyri; the parietal lobe, including the superior and inferior parietal lobules; the occipital lobe, containing the striate cortex; the hippocampus, sampled at the level of the dentate gyrus; the basal ganglia, including the caudate, putamen, and globus pallidus; the thalamus, and subthalamic nucleus; the midbrain, at the level of the red nucleus; and the cerebellum, at the level of the dentate nucleus. Immunohistochemical staining for α-synuclein (MA1-90342; Thermo Scientific; 1:1500), amyloid-β (M0872; Dako; 1:100), phosphorylated tau (MN1020; Thermo Scientific; 1:1,200) and non-phosphorylated TDP-43 (2E2-D3, H00023435-M01, Abnova, 1:500) was performed using automated staining platforms from Menarini Diagnostics or Ventana (Roche). Biotinylated secondary antibodies were used with horseradish peroxidase-conjugated streptavidin complex and diaminobenzidine as the chromogen, with haematoxylin counterstaining. Appropriate positive and negative controls were included in each staining run. Pathological diagnoses were confirmed by neuropathologists experienced in neurodegenerative diseases using relevant consensus diagnostic and staging criteria including those developed for PSP by Kovacs and colleagues.^5,35–40^

### Quantitative digital neuropathology

Immunohistochemistry-stained whole slides from eight brain regions (anterior frontal, posterior frontal, temporal and parietal lobes, hippocampus sampled at the level of the dentate gyrus, basal ganglia, midbrain at the level of the red nucleus, and the cerebellum at the level of the dentate nucleus) were scanned on a Hamamatsu NanoZoomer S360 (26×76 mm slides; 40× objective; JPEG compression quality 70; pixel size 0.23 µm/pixel) or Hamamatsu NanoZoomer S60 (double 52×76 mm slides; 40× objective; JPEG compression quality 65; pixel size 0.22 µm/pixel) slide scanner (Fig. 1). The subthalamic nucleus, a characteristic site of PSP pathology, was excluded from quantitative analysis due to the difficulty of accurately delineating its boundaries in the context of the significant degree of atrophy commonly observed in this region. Forty-one anatomical regions of interest (ROIs) (Supplementary Table 1) were manually annotated on the digitised sections using a web-based whole slide image (WSI) viewer (NZConnect, Hamamatsu). The annotations were exported and downloaded as NDPA files using a custom Python script and imported into QuPath v0.5.1^41^ for image analysis using a custom Groovy script. Stain vectors for colour deconvolution were calculated from a composite image consisting of 1000×1000-pixel squares derived from 270 WSIs stained for phosphorylated tau, with 1% of extreme pixels ignored. After applying the stain vectors onto each WSI, a threshold for positive phosphorylated tau staining was calculated using the Otsu^42^ automatic thresholding method via an adaptation of FIJI/ImageJ’s implementation for each brain region annotation. The automatically calculated threshold values were then used to measure the area of phosphorylated tau staining within each annotated ROI. The percentage of positive DAB staining within each brain region was calculated by: %Tau staining area = (area of phosphorylated tau staining/area of brain region annotation) ×100. We created composite values for grey matter (GM) and white matter (WM) across cortical, limbic, basal ganglia, midbrain, and cerebellar regions for summary statistics (Supplementary Table 2).

**Figure 1.**
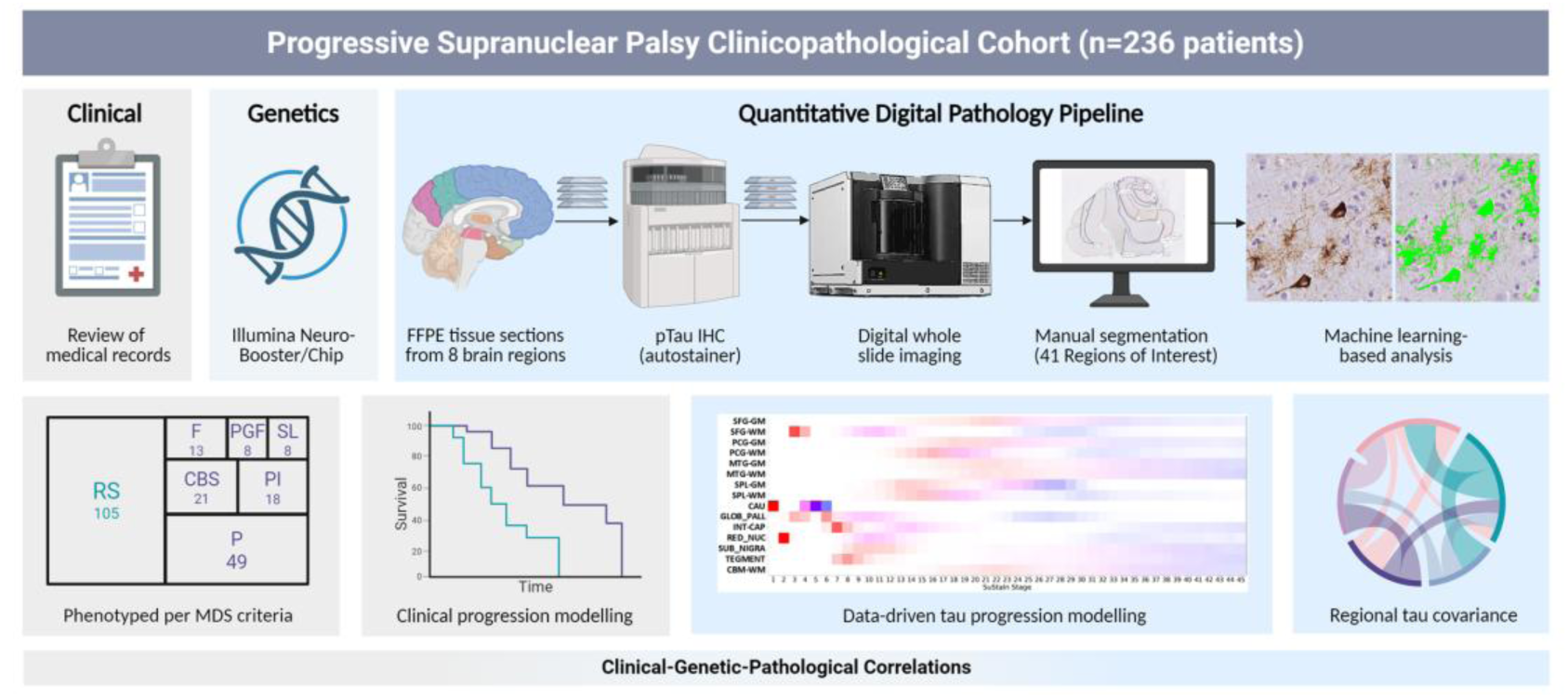
Clinical, genetic and quantitative digital pathology pipeline. Schematic representation of clinicogenetic and quantitative digital pathology pipeline. FFPE: formalin fixed paraffin embedded; pTau: phosphorylated tau; IHC: immunohistochemistry.

### Data-driven modelling

The Subtype and Stage Inference (SuStaIn) algorithm combines progression modelling with clustering to facilitate the identification of subgroups with distinct progression patterns from cross-sectional data.^15^ Depending on the data type, subgroup progression patterns can be modelled using different approaches; in this study we used the piecewise linear z-score trajectory approach for continuous data. We applied SuStaIn to quantitative tau pathology data from 15 ROIs (Supplementary Table 1), selected to include both the canonical brain regions typically affected by PSP pathology and based on quantitative tau pathology data, those regions most effective at distinguishing PSP clinical phenotypes. Neurologically healthy aged controls rarely exhibit incidental PSP tau pathology but often show incidental AD tau pathology, making them an unsuitable reference population for regional quantitative tau pathology data aimed at characterising PSP. Therefore, to meet the SuStaIn framework’s requirement for z-score normalisation and in the absence of appropriate control tissue, we generated synthetic control profiles instead of using data from neurotypical patients. A total of 27 synthetic control subjects were constructed to reflect minimal to absent tau pathology. For 25 synthetic control profiles, values were generated for each brain region simulating 0–5% of the true pathological burden with an approximately normal distribution. To test model robustness, two additional synthetic outlier profiles with moderate pathology levels (∼30–50%) were generated and included in the analysis.

The SuStaIn algorithm optimises the assignment of individuals to subtypes and the sequence in which ROIs reach different z-scores within each subtype using a data likelihood function. Model uncertainty was assessed with 100,000 Markov Chain Monte Carlo (MCMC) iterations, and the expectation-maximisation process optimised the subtype sequences from 25 random starting points to find the maximum likelihood solution. We evaluated models comprising two to six SuStaIn subtypes. To determine the optimal number of subtypes, we performed 10-fold cross-validation across models and calculated the cross-validation information criterion (CVIC), which evaluates the balance of model accuracy against model complexity.^15^ We also inspected the test-set log-likelihoods plots, which assesses the generalisability of the model, and the MCMC log-likelihood traces and histograms, which evaluate model fit with increasing subtypes.

### Statistical analysis

To assess differences in demographic, clinical, genetic, and neuropathological features across PSP clinical phenotypes we used Pearson’s chi-square or Fisher’s exact test for categorical variables, as appropriate; *P*-values were estimated using Monte Carlo simulation for sparse contingency tables. For categorical variables with more than two levels, we conducted post-hoc pairwise Fisher’s exact tests between all phenotype pairs using the same simulation approach with Bonferroni correction for multiple comparisons. For continuous variables we used one-way ANOVA followed by pairwise comparisons using estimated marginal means with Bonferroni correction. For continuous variables that violated parametric assumptions, including survival and latencies to individual clinical milestones we used the Kruskal–Wallis test with post hoc Dunn’s tests and Bonferroni correction. To assess clinical predictors of survival, we conducted Cox proportional hazards regression using the survival package in R. Time interaction models were used where proportional hazard assumptions were violated, and the analysis was stratified by phenotype to allow the baseline hazard to differ across its levels. Generalised linear models with a gamma distribution and log link were applied to assess the relationship between tau pathology burden and survival adjusting for age at symptom onset, phenotype, sex, and National Institute on Aging-Alzheimer’s Association ABC (NIA-ABC) score, a staging system that integrates amyloid-β pathology regional burden (A), neurofibrillary tangle stage (B), and neuritic plaque density (C) to assess the overall severity of AD pathology.^43^ Differences in regional tau pathology burden across PSP clinical phenotypes were assessed using the Kruskal–Wallis test, followed by pairwise Wilcoxon rank-sum tests where significant, with Bonferroni correction applied within each region; this was conducted both on raw values and on residuals from linear regression adjusting for age, sex, and NIA-ABC score. To evaluate regional covariance in tau pathology, linear regression was performed using age, sex and NIA-ABC score as covariates, and then pairwise Spearman correlations were computed on the residuals. Strong correlations were defined as those with Bonferroni adjusted *P* < 0.05 and Spearman’s ρ > 0.5. Regional tau pathology burden was compared across sex, *MAPT* H2 haplotype, *APOE* ε4, *TRIM11* (A), and *SLC2A13* (A) status using Wilcoxon rank-sum tests, with Benjamini-Hochberg correction for multiple comparisons across regions.

To assess differences in regional tau pathology by SuStaIn subtype, Kruskal–Wallis tests were applied to measurements across SuStaIn subtypes 1–3. Dunn’s post hoc tests were used for pairwise comparisons, with Bonferroni correction; Bonferroni was applied for comparisons across clinical subtypes to prioritise specificity, while Benjamini-Hochberg was used for comparisons across brain regions to enhance sensitivity given the large number of brain regions. To assess the relationship between SuStaIn stage and survival, we residualised both SuStaIn stage and disease duration using linear models that controlled for age at onset and sex and then computed Spearman rank correlations between the residuals for each SuStaIn stage. The association between SuStaIn stage and Kovacs’ pathological stages was performed using Spearman’s correlation. Cases with missing values were excluded listwise for each analysis, based on the specific variables required. All *P*-values were 2-tailed. Statistical analysis was performed using SPSS version 29 (IBM) and R (4.4.2), and graphs were generated using R. A large language model (ChatGPT, OpenAI) was used to assist in generating R code for statistical analyses and data visualisation. All code, statistical tests, and outputs were manually reviewed and validated by the authors to ensure methodological appropriateness and accuracy of results.

### Data availability

Anonymised clinical and quantitative tau pathology data may be shared upon reasonable request by any qualified investigator for purposes of replicating procedures and results, if in compliance with ethical approval.

## Results

### Demographic, clinicopathological and genetic characteristics of the cohort

We identified 241 pathologically confirmed PSP patients. Five patients were excluded due to insufficient clinical data. The mean ages at symptom onset and death were 67.8±7.5 and 75.8±7.4 years, respectively. Thirteen patients could not be assigned a phenotype due to inadequate documentation of eye movement examination at 3 years, and one patient had a primary lateral sclerosis phenotype, which was not classifiable per MDS criteria. Demographic and clinical data for the remaining 222 patients grouped by MDS phenotype at 3 years from symptom onset are shown in Table 1. Overall, 63.6% of patients in the cohort were male, but 66.7% of PSP-corticobasal syndrome (PSP-CBS) and postural instability (PSP-PI) phenotypes were female. PSP-frontal (PSP-F) and PSP-speech/language (PSP-SL) patients had the earliest and latest ages of symptom onset and death, respectively. Levodopa responsiveness in PSP with predominant parkinsonism (PSP-P) was twice that of PSP-RS, but sustained responsiveness beyond two years did not differ across phenotypes. Genetic data was available for 193 patients. *TRIM11* (A) was less frequent in patients with PSP-P and PSP with pure gait freezing (PSP-PGF) clinical phenotypes, but the difference was not significant and there was no difference in the frequency of *MAPT* H2 haplotype, *APOE* ε4, or *SLC2A13* (A) across clinical phenotypes. Mean ages at symptom onset and death were 6.1 and 5.7 years later in *MAPT* H2 carriers compared to *MAPT* H1/H1 carriers. There were no significant differences in the semi-quantitative staging of AD or Lewy body pathologies across PSP phenotypes as assessed by Thal phases, Braak and Braak stages for tau pathology and Braak staging for Lewy pathology. Limbic TDP-43 pathology was present in 87.5% of PSP-SL cases, considerably more than all other clinical phenotypes.

**Table 1.**
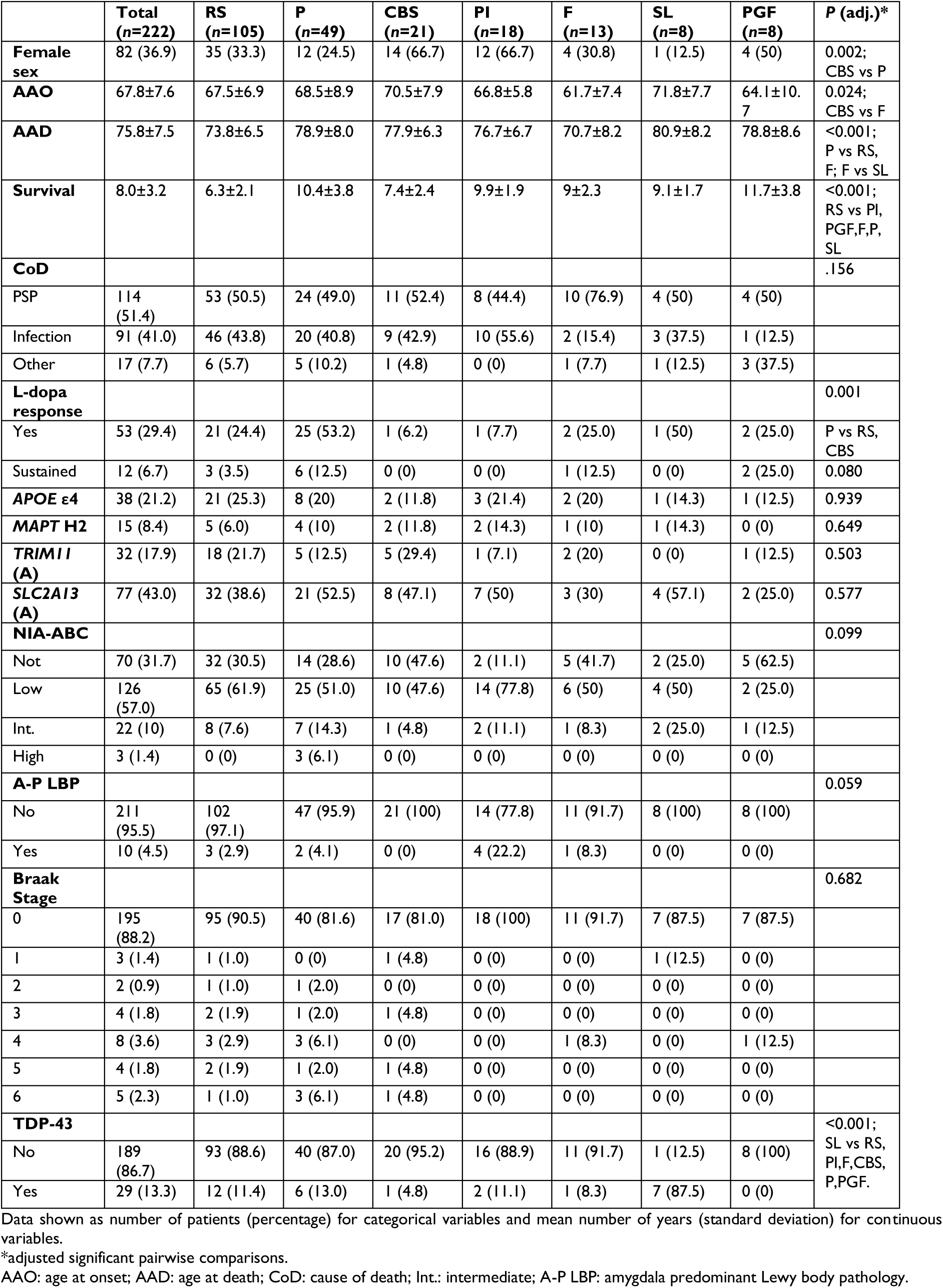
Demographic, clinical and pathological characteristics by PSP subtype.

|  | **Total**<br>**(n=222)** | **RS**<br>**(n=105)** | **P**<br>**(n=49)** | **CBS**<br>**(n=21)** | **PI**<br>**(n=18)** | **F**<br>**(n=13)** | **SL**<br>**(n=8)** | **PGF**<br>**(n=8)** | **P (adj.)\*** |
| --- | --- | --- | --- | --- | --- | --- | --- | --- | --- |
| **Female sex** | 82 (36.9) | 35 (33.3) | 12 (24.5) | 14 (66.7) | 12 (66.7) | 4 (30.8) | 1 (12.5) | 4 (50) | 0.002;<br>CBS vs P |
| **AAO** | 67.8±7.6 | 67.5±6.9 | 68.5±8.9 | 70.5±7.9 | 66.8±5.8 | 61.7±7.4 | 71.8±7.7 | 64.1±10.7 | 0.024;<br>CBS vs F |
| **AAD** | 75.8±7.5 | 73.8±6.5 | 78.9±8.0 | 77.9±6.3 | 76.7±6.7 | 70.7±8.2 | 80.9±8.2 | 78.8±8.6 | <0.001;<br>P vs RS,<br>F; F vs SL |
| **Survival** | 8.0±3.2 | 6.3±2.1 | 10.4±3.8 | 7.4±2.4 | 9.9±1.9 | 9±2.3 | 9.1±1.7 | 11.7±3.8 | <0.001;<br>RS vs PI,<br>PGF,F,P,<br>SL |
| **CoD** |  |  |  |  |  |  |  |  | .156 |
| PSP | 114 (51.4) | 53 (50.5) | 24 (49.0) | 11 (52.4) | 8 (44.4) | 10 (76.9) | 4 (50) | 4 (50) |  |
| Infection | 91 (41.0) | 46 (43.8) | 20 (40.8) | 9 (42.9) | 10 (55.6) | 2 (15.4) | 3 (37.5) | 1 (12.5) |  |
| Other | 17 (7.7) | 6 (5.7) | 5 (10.2) | 1 (4.8) | 0 (0) | 1 (7.7) | 1 (12.5) | 3 (37.5) |  |
| **L-dopa response** |  |  |  |  |  |  |  |  | 0.001 |
| Yes | 53 (29.4) | 21 (24.4) | 25 (53.2) | 1 (6.2) | 1 (7.7) | 2 (25.0) | 1 (50) | 2 (25.0) | P vs RS,<br>CBS |
| Sustained | 12 (6.7) | 3 (3.5) | 6 (12.5) | 0 (0) | 0 (0) | 1 (12.5) | 0 (0) | 2 (25.0) | 0.080 |
| **APOE ε4** | 38 (21.2) | 21 (25.3) | 8 (20) | 2 (11.8) | 3 (21.4) | 2 (20) | 1 (14.3) | 1 (12.5) | 0.939 |
| **MAPT H2** | 15 (8.4) | 5 (6.0) | 4 (10) | 2 (11.8) | 2 (14.3) | 1 (10) | 1 (14.3) | 0 (0) | 0.649 |
| **TRIM11 (A)** | 32 (17.9) | 18 (21.7) | 5 (12.5) | 5 (29.4) | 1 (7.1) | 2 (20) | 0 (0) | 1 (12.5) | 0.503 |
| **SLC2A13 (A)** | 77 (43.0) | 32 (38.6) | 21 (52.5) | 8 (47.1) | 7 (50) | 3 (30) | 4 (57.1) | 2 (25.0) | 0.577 |
| **NIA-ABC** |  |  |  |  |  |  |  |  | 0.099 |
| Not | 70 (31.7) | 32 (30.5) | 14 (28.6) | 10 (47.6) | 2 (11.1) | 5 (41.7) | 2 (25.0) | 5 (62.5) |  |
| Low | 126 (57.0) | 65 (61.9) | 25 (51.0) | 10 (47.6) | 14 (77.8) | 6 (50) | 4 (50) | 2 (25.0) |  |
| Int. | 22 (10) | 8 (7.6) | 7 (14.3) | 1 (4.8) | 2 (11.1) | 1 (8.3) | 2 (25.0) | 1 (12.5) |  |
| High | 3 (1.4) | 0 (0) | 3 (6.1) | 0 (0) | 0 (0) | 0 (0) | 0 (0) | 0 (0) |  |
| **A-P LBP** |  |  |  |  |  |  |  |  | 0.059 |
| No | 211 (95.5) | 102 (97.1) | 47 (95.9) | 21 (100) | 14 (77.8) | 11 (91.7) | 8 (100) | 8 (100) |  |
| Yes | 10 (4.5) | 3 (2.9) | 2 (4.1) | 0 (0) | 4 (22.2) | 1 (8.3) | 0 (0) | 0 (0) |  |
| **Braak Stage** |  |  |  |  |  |  |  |  | 0.682 |
| 0 | 195 (88.2) | 95 (90.5) | 40 (81.6) | 17 (81.0) | 18 (100) | 11 (91.7) | 7 (87.5) | 7 (87.5) |  |
| 1 | 3 (1.4) | 1 (1.0) | 0 (0) | 1 (4.8) | 0 (0) | 0 (0) | 1 (12.5) | 0 (0) |  |
| 2 | 2 (0.9) | 1 (1.0) | 1 (2.0) | 0 (0) | 0 (0) | 0 (0) | 0 (0) | 0 (0) |  |
| 3 | 4 (1.8) | 2 (1.9) | 1 (2.0) | 1 (4.8) | 0 (0) | 0 (0) | 0 (0) | 0 (0) |  |
| 4 | 8 (3.6) | 3 (2.9) | 3 (6.1) | 0 (0) | 0 (0) | 1 (8.3) | 0 (0) | 1 (12.5) |  |
| 5 | 4 (1.8) | 2 (1.9) | 1 (2.0) | 1 (4.8) | 0 (0) | 0 (0) | 0 (0) | 0 (0) |  |
| 6 | 5 (2.3) | 1 (1.0) | 3 (6.1) | 1 (4.8) | 0 (0) | 0 (0) | 0 (0) | 0 (0) |  |
| **TDP-43** |  |  |  |  |  |  |  |  | <0.001;<br>SL vs RS,<br>PI,F,CBS,<br>P,PGF. |
| No | 189 (86.7) | 93 (88.6) | 40 (87.0) | 20 (95.2) | 16 (88.9) | 11 (91.7) | 1 (12.5) | 8 (100) |  |
| Yes | 29 (13.3) | 12 (11.4) | 6 (13.0) | 1 (4.8) | 2 (11.1) | 1 (8.3) | 7 (87.5) | 0 (0) |  |

Data shown as number of patients (percentage) for categorical variables and mean number of years (standard deviation) for continuous variables.
\*adjusted significant pairwise comparisons.
AAO: age at onset; AAD: age at death; CoD: cause of death; Int.: intermediate; A-P LBP: amygdala predominant Lewy body pathology.

### Early clinical and pathological predictors of survival

Overall, the mean survival from symptom onset was 8±3.2 years. Mean survival and latencies to clinical milestones between PSP phenotypes are shown in Fig. 2A, and significant differences in the timing of clinical milestones and survival between phenotypes are shown in Fig. 2B. Cause of death did not differ by clinical phenotype (Table 1). No major clinical milestones differed by sex. After adjusting for onset age, clinical phenotype and sex, several clinical features present within the first three years of symptom onset predicted shorter survival, including vertical gaze palsy, axial and limb rigidity, dysexecutive symptoms, bradyphrenia, apraxia, bulbar symptoms, constipation, urinary symptoms, clinical REM sleep behaviour disorder, insomnia and visual hallucinations (Fig. 2C). Importantly, there was no significant difference in the frequency of Lewy body pathology among patients who experienced constipation, clinical REM sleep behaviour disorder, or visual hallucinations within the first three years after symptom onset. Several other early clinical features were also associated with reduced survival in univariate analyses (Fig. 2C). Regional quantitative tau pathology data were available for 235 cases; missing data for individual regions are detailed in Supplementary Table 1. Adjusting for onset age, phenotype, sex and NIA-ABC score, survival was inversely associated with quantitative tau pathology burden in the cerebellar dentate nucleus, several midbrain and basal ganglia GM areas as well as WM in the centrum semiovale, precentral gyrus and superior parietal lobule (Fig. 2D, Supplementary Table 3). Among the covariates, age at symptom onset was also significantly associated with survival.

**Figure 2.**
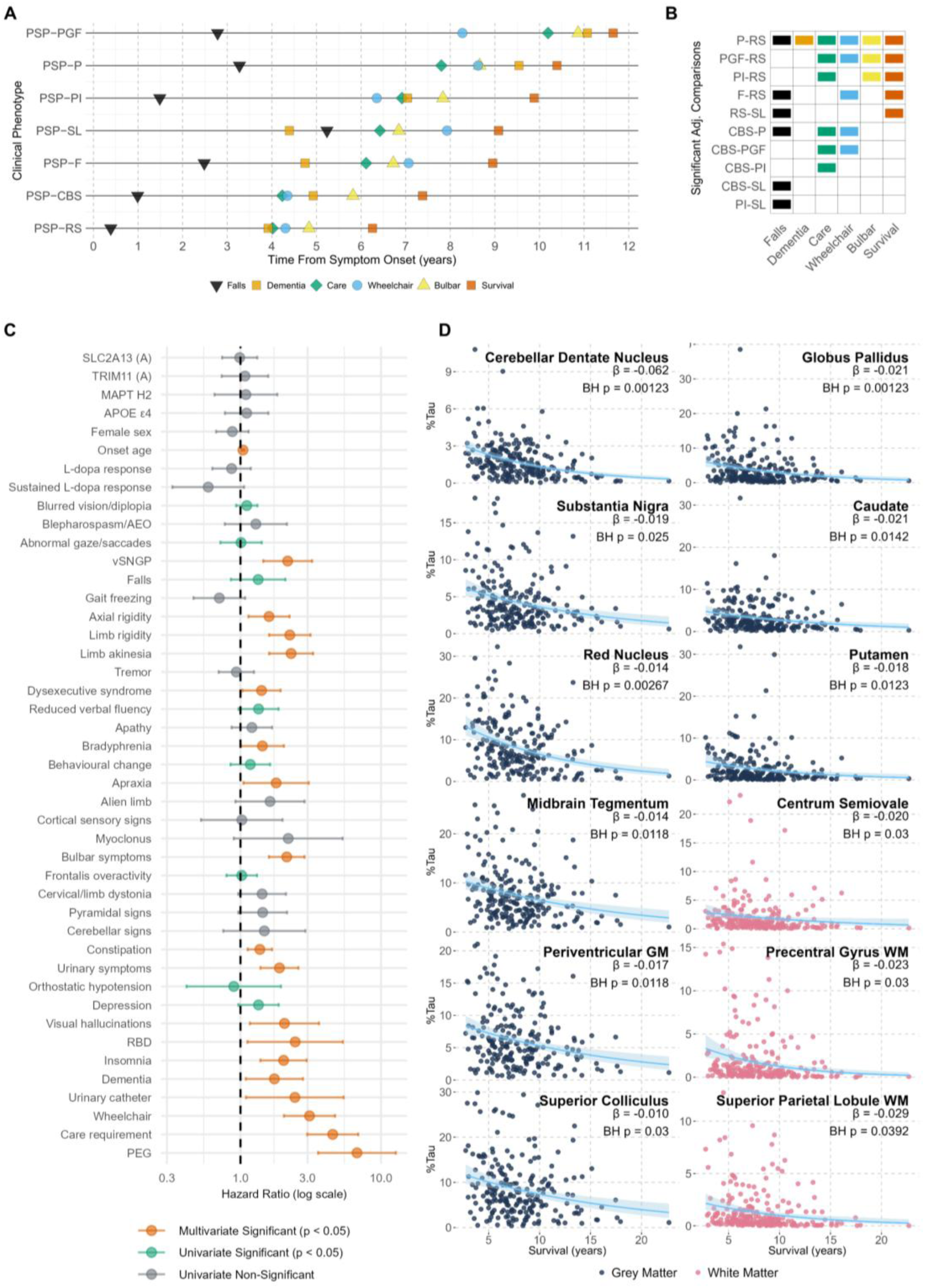
Clinical milestones by phenotype, and clinical and pathological predictors of survival. **(A)** The line plot shows the mean time from symptom onset to key clinical milestones and survival for each PSP phenotype. **(B)** The tile plot displays significant pairwise differences in milestone timing and survival between phenotypes; coloured tiles indicate milestones with significant differences for each phenotype comparison (Bonferroni-corrected *P* < 0.05). **(C)** The Forest plot shows hazard ratios (HRs) and 95% confidence intervals for overall survival across multiple clinical and genetic variables. Variables not reaching significance in univariate analyses are shown in grey; those significant only in univariate analyses are shown in green; and variables significant in adjusted multivariate models are shown in orange. Multivariate models were adjusted for sex, age at onset, and baseline phenotype with time-interaction terms included if proportional hazards assumptions were violated. **(D)** Scatter plots show the relationship between tau pathology burden (%Tau) in individual brain regions and survival adjusting for onset age, phenotype, sex, and NIA-ABC score, with each point representing an individual patient. Each panel displays a fitted Gamma regression line with 95% confidence interval shading. The regions shown were statistically significant after Benjamini–Hochberg correction. β coefficients and adjusted *P*-values are annotated for each model. vSNGP: vertical supranuclear gaze palsy; RBD: REM sleep behaviour disorder.

### Clinical and genetic determinants of regional tau pathology

We identified anatomically congruent differences in quantitative tau pathology burden between clinical phenotypes, including higher GM and WM pathology in the superior and middle temporal gyri of PSP-SL patients, the precentral gyrus and superior parietal lobule of PSP-CBS, and in precentral gyrus and several subcortical regions in PSP-RS compared to PSP-P (Fig. 3A and B). Male patients had nominally higher tau pathology in the superior colliculus, caudate, and hippocampal CA3 subregion, although these differences were not significant after correcting for multiple comparisons (Fig. 4). Restricting the analysis to PSP-RS patients, tau pathology in males was even greater in the midbrain, with additional nominal differences in the red nucleus, midbrain tegmentum and periventricular GM (Supplementary Fig. 1). In patients with PSP-CBS phenotype, males had nominally higher tau in precentral gyrus GM and WM (Supplementary Fig. 1). *MAPT* H2 haplotype carriers had significantly higher tau pathology than H1 carriers in several cortical and limbic regions, centrum semiovale WM, and cerebellar dentate nucleus (Fig. 4). *TRIM11* (A) carriers had nominally lower tau pathology in the hippocampal dentate gyrus, entorhinal and transentorhinal cortices and parahippocampal gyrus WM. Regional tau pathology did not vary by *APOE* ε4 or *SLC2A13* (A) status (Fig. 4). After adjusting for sex, phenotype and NIA-ABC score, age at symptom onset was positively correlated with quantitative tau pathology burden in limbic GM and WM and inversely associated with midbrain GM and centrum semiovale WM. Age at death was positively correlated with limbic GM and WM tau pathology and inversely correlated with midbrain GM and centrum semiovale and cerebellar WM tau pathology burden (Supplementary Table 4).

**Figure 3.**
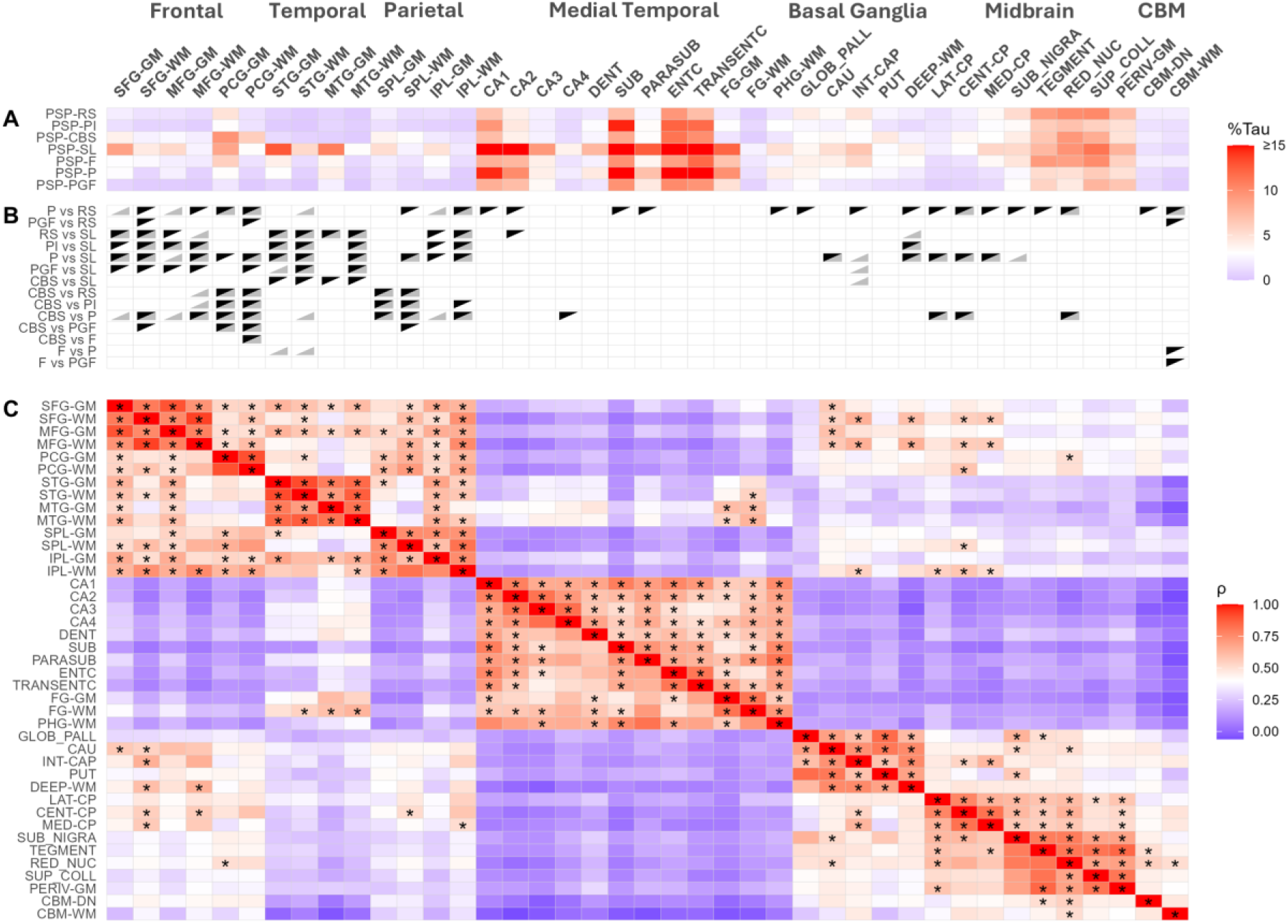
Regional tau pathology burden by clinical phenotype and regional covariance. **(A)** The upper heatmap shows mean tau pathology burden (% Tau) in 41 individual brain regions for each clinical phenotype. **(B)** The tile plot shows brain regions where there are significant pairwise differences in tau pathology burden between phenotypes; within each tile, a black upper triangle indicates a statistically significant pairwise difference (*P* < 0.05, Bonferroni-corrected) in the unadjusted analysis and a grey lower triangle indicates significance in the analysis adjusted for age, sex, and NIA-ABC score. **(C)** The correlation heatmap shows pairwise Spearman correlations between residualised tau pathology values adjusted for age, sex, and NIA-ABC score; asterisks mark correlations with ρ >0.5 that were significant after Bonferroni correction for multiple testing. CBM: cerebellum.

**Figure 4.**
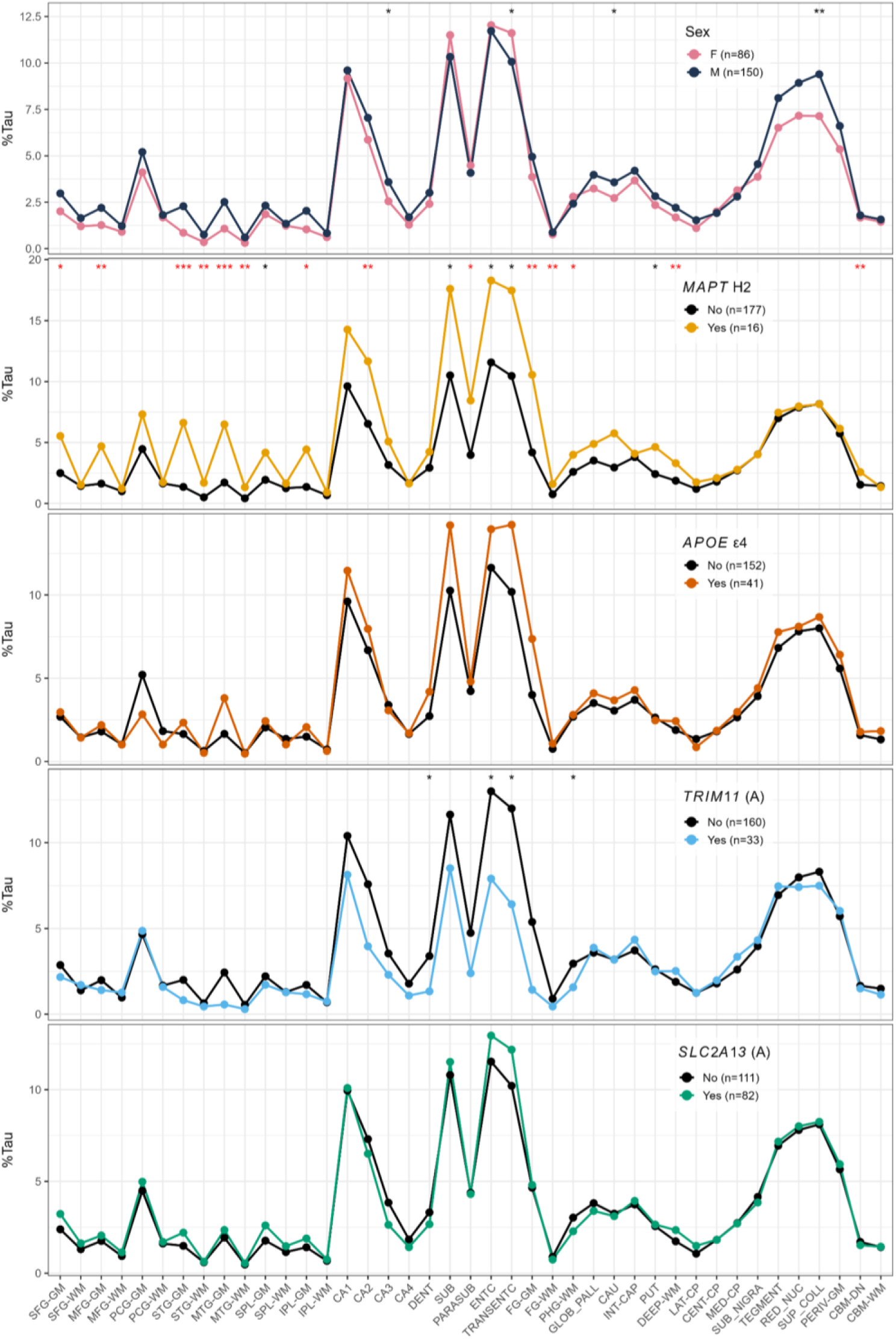
Regional tau pathology by sex and genotype. The line plots show mean tau pathology burden (% Tau) across 41 brain regions for each category of sex, *MAPT* H2, *APOE* ε4, *TRIM11* (A), and *SLC2A13* (A) in all PSP cases. Error bars have been omitted for clarity. Asterisks indicate statistically significant differences between groups in individual regions (*P* < 0.05 = *, *P* < 0.01 = **, *P* < 0.001 = ***). Red significance symbols denote regions that remain significant following Benjamini–Hochberg correction for multiple comparisons. Sample sizes (*n*) for each group are indicated in the legends.

### Regional covariance in quantitative tau pathology

In general, tau pathology in adjacent ROIs correlated strongly with each other (Fig. 3C). Tau pathology burden in the middle temporal gyrus correlated positively with that of fusiform gyrus GM and WM. Significant correlations in tau pathology burden were also observed between ROIs in anatomically connected but spatially distributed brain regions, including between superior and middle frontal gyri WM and centrum semiovale, internal capsule and cerebral peduncle WM. There were also strong significant correlations between superior frontal gyrus GM and WM pathology and that of the caudate nucleus, while precentral gyrus GM tau pathology was correlated with red nucleus pathology. Tau pathology in both substantia nigra and midbrain tegmentum correlated with the pathology load in the caudate. Cerebellar dentate nucleus and white matter tau pathology burden also correlated with that of the red nucleus.

### Spatiotemporal trajectories of PSP tau pathology progression

Applying SuStaIn to quantitative tau pathology data from all PSP patients yielded three subtypes, including one containing only patients with intermediate/high NIA-ABC scores or significant primary ageing-related tauopathy (Supplementary Fig. 3). To minimise the effect of AD co-pathology, we repeated SuStaIn after excluding these 12 patients and identified three SuStaIn subtypes for ‘pure’ PSP (Fig. 5A-C). Although the CVIC, reached its minimum value in the five-subtype model (Fig. 5D), indicating improved fit with up to five subtypes, the test-set log-likelihoods plateaued after three subtypes (Fig. 5E), indicating limited generalisability beyond three subtypes. In addition, the MCMC log-likelihood traces and histograms for models with more than three subtypes exhibited substantial overlap (Fig. 5F), suggesting that additional subtypes did not meaningfully improve model fit. In contrast, subtypes 1 to 3 showed clear, incremental gains in log-likelihood. The choice of three subtypes was further supported by inspection of the positional variance diagrams (PVDs), which showed low positional uncertainty and distinct, anatomically plausible progression patterns in the three-subtype model (Fig. 5A-C). In contrast, the four- and five-subtype models introduced more fragmented or overlapping trajectories with increased uncertainty.

**Figure 5.**
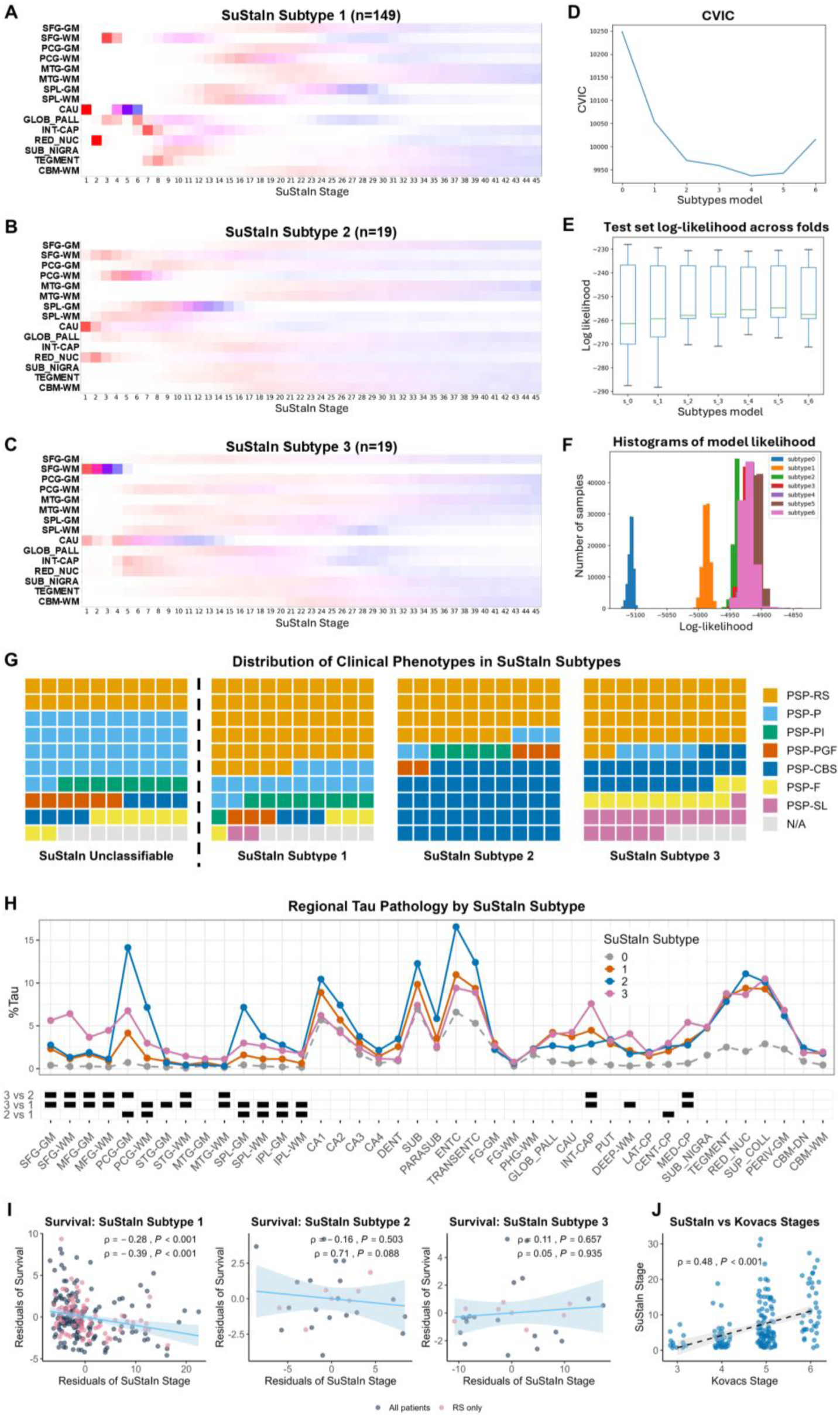
Subtype and Stage Inference modelling of ‘pure’ PSP pathology. **(A-C)** Positional variance diagrams show the progression patterns of regional tau pathology inferred by the SuStaIn model for each of the three data- driven subtypes. Each row corresponds to a specific brain region, and each column to a SuStaIn stage; colour intensity reflects the probability that a given brain region is affected at a specific stage, with red indicating higher probability, blue lower probability, and white indicating low or no probability. **(D)** The line plot shows cross-validated information criterion (CVIC) scores for SuStaIn models with increasing numbers of subtypes; lower CVIC values indicate better model fit. **(E)** The boxplots of test-set likelihoods across cross-validation folds reflect model generalisability. **(F)** The histogram displays model likelihoods, summarising the distribution of likelihood values across the dataset. **(G)** The waffle plots show the proportional distribution of PSP clinical phenotypes within each SuStaIn subtype and unclassifiable cases, with each square representing 1% of the corresponding subtype. **(H)** The lower line plot displays mean regional tau pathology burden across individual brain regions for SuStaIn subtypes 1-3 and unclassifiable cases (subtype 0), and the corresponding tile plot highlights regions with significant pairwise group differences in tau pathology burden; black tiles indicate regions with Benjamini– Hochberg adjusted *P* < 0.05. **(I)** The scatter plots show the relationship between SuStaIn stage and survival across SuStaIn subtypes 1-3; points represent residuals from linear models, and lines (blue) show fitted regressions with 95% confidence intervals (shaded) for analyses including all patients (navy) and analyses restricted to PSP-RS patients only (coral). Correlation coefficients (*ρ*) and associated *P*-values are annotated. **(J)** The final panel compares machine learning derived SuStaIn stages with semi-quantitatively assigned Kovacs’ stages, with black dashed regression line and grey confidence band; the correlation coefficient (*ρ*) and associated *P*-value are annotated.

SuStaIn-defined subtype 1 shows progression from subcortical GM nuclei to superior frontal WM, followed by involvement of other midbrain structures and, subsequently, other cortical regions (Fig. 5A). Subtype 2 and subtype 3 show simultaneous early involvement of the caudate nucleus and superior frontal gyrus WM (Fig. 5B and C). Subtype 2 is distinguished by early involvement of the red nucleus, and an emphasis on more posterior precentral gyrus and superior parietal lobule WM followed by involvement of corresponding cortical GM areas (Fig. 5B). In Subtype 3 the emphasis is more anterior in the superior frontal WM with subsequent involvement of precentral gyrus, middle temporal gyrus and superior parietal lobule WM, again followed by involvement of corresponding cortical GM areas (Fig. 5C). Subcortical phenotypes (PSP-RS, PSP-PI, PSP-P and PSP-PGF cases) combined comprised 83.2% of subtype 1 patients compared to 52.6% and 47.4% of subtype 2 and subtype 3 respectively (Fig. 5G). Conversely, cortical phenotypes (PSP-CBS, PSP-SL, and PSP-F cases) comprised a considerably greater proportion of subtype 2 and subtype 3 patients (47.4% each) compared to subtype 1 (9.4%). Specifically, 85% of patients classified as RS and 93% of both PSP-P and PSP-PI patients were included in subtype 1, whilst 56% of those with either PSP-CBS, PSP-SL or PSP-F were categorised in subtype 2 or 3. There were no differences in sex, genetic factors, age at symptom onset, or disease duration between SuStaIn subtypes (Supplementary Table 5). Compared to subtype 1, tau pathology was higher in several regions of subtype 3 patients including the superior and middle frontal gyri WM and GM, centrum semiovale, internal capsule and medial cerebral peduncle (Fig. 5H). Tau pathology was greater in precentral gyrus and superior and inferior parietal lobule GM and WM in subtype 2 (Fig. 5H). Thirty-six patients were not classifiable by SuStaIn (subtype 0) and the mean tau pathology load was considerably lower in almost all regions in these patients (Fig. 5H). Almost half of the unclassifiable patients had a PSP-P phenotype (*n*=15), followed by PSP-RS (*n*=7), PSP-PI (*n*=3), PSP-CBS (*n*=3), PSP-F (*n*=3), PSP-PGF (*n*=2), no MDS phenotype assigned (*n*=3). Adjusting for sex and onset age, SuStaIn stage was inversely correlated with survival in subtype 1, and the strength of the association increased when restricting the analysis to PSP-RS cases (Fig. 5I). There were no significant correlations between SuStaIn stage and survival in subtypes 2 and 3 (Fig. 5I). Semi-quantitative Kovacs stages^5^ corresponded well to both SuStaIn stage (Fig. 5J) and quantitative measurements of regional tau pathology (Fig. 6).

**Figure 6.**
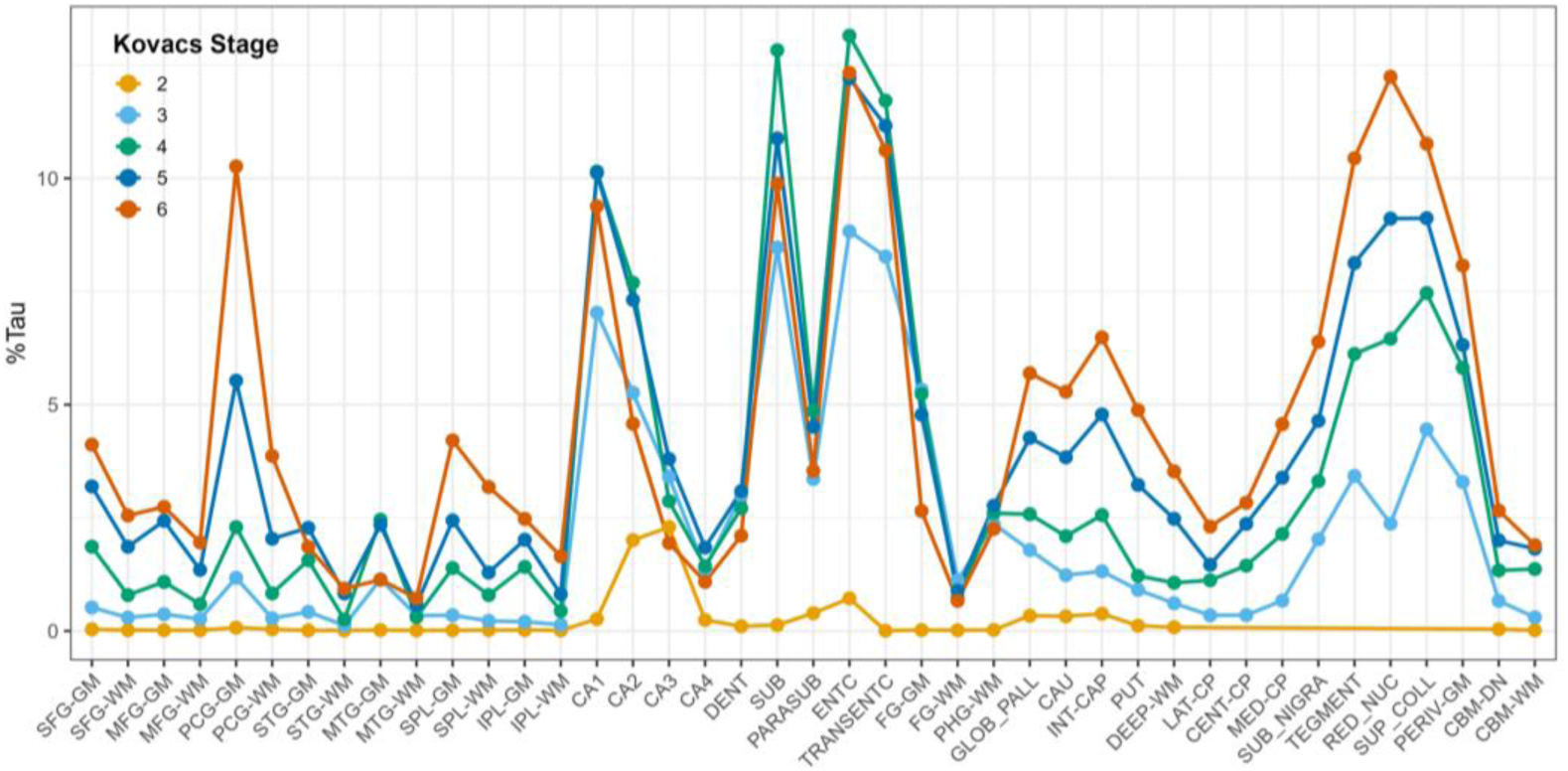
Mean regional tau pathology burden across semi-quantitatively defined PSP stages. Each line shows mean quantitative tau pathology burden (%Tau) in 41 brain regions across each semi-quantitatively scored PSP stage.

## Discussion

Using standardised clinical criteria and automated digital quantitative pathology methods, this comprehensive clinicopathological study has elucidated: 1) the longitudinal clinical course and clinical features that predict survival in a large cohort of patients with PSP; 2) region specific differences in tau pathology burden associated with phenotypic diversity and clinical progression; 3) specific contributions of sex, genotype and co-pathology toward clinical phenotype and regional tau pathology burden; and 4) spatiotemporal patterns of tau pathology covariance and progression.

The overall survival observed in our cohort was consistent with previous studies reporting mean disease durations of 7-8 years for PSP overall and 5-7 years for PSP-RS.^12,13,21,44–48^ Comparing non-RS phenotype-specific survival across studies is more challenging because of the lack of standardised diagnostic criteria before 2017, compounded by the fact that PSP clinical phenotype is not necessarily stable over time^49^. Also consistent with previous clinicopathological studies, features associated with reduced survival included older age at symptom onset,^21,48,50–52^ dysautonomia,^53^ urinary incontinence,^46^ early falls^46,50,52,54^ and supranuclear gaze palsy,^54–56^ bulbar dysfunction,^46,47,50,51^ and cognitive symptoms.^47,51,52,57^ Our results also support the clinical observation in a small number of patients that sleep disorders and possibly visual hallucinations are predictive of shorter survival in patients with PSP.^58^

We identified several phenotype-specific regional tau pathology signatures that support and extend previous observations that clinical phenotype corresponds to the neuroanatomical distribution of tau pathology,^5–9,59^ although direct comparisons are again challenging because of non-standardised clinical phenotyping and the practice of combining phenotypes to increase statistical power.^60^ As anticipated from previous studies,^5,9,61^ PSP-RS patients exhibited a significantly higher tau pathology burden across several neocortical, basal ganglia, midbrain and cerebellar structures compared to PSP-P, and PSP-PGF patients had the lowest tau pathology across neocortical regions and also cerebellar white matter. Tau pathology burden in PSP-CBS patients was greatest in the precentral gyrus and superior parietal lobule, while the heaviest pathology in PSP-SL was in the superior and middle temporal gyri GM. However, precentral gyrus WM rather than GM was most discriminatory for PSP-CBS, while tau pathology load in middle temporal WM was more specific for the PSP-SL phenotype than corresponding GM pathology burden. This distribution of pathology is consistent with widespread patterns of white matter tract degeneration identified on diffusion weighted MRI of patients with PSP-SL.^62^ As in previous clinicopathological work using MDS clinical criteria,^5^ PSP-F patients did not have a distinct cortical tau pathology signature, which may reflect limitations in the anatomical regions of the frontal lobe available for analysis, the specificity of clinical criteria, or pathomechanisms relating to network disruption or cell-autonomous vulnerability.

Regarding regional tau pathology burden and progression, we identified 12 specific brain regions (cerebellar dentate nucleus, substantia nigra, red nucleus, midbrain tegmentum, including periventricular grey matter and superior colliculus, globus pallidus, caudate, putamen, centrum semiovale, precentral gyrus and superior parietal lobule WM) where tau pathology burden is associated with survival, including the dentate nucleus, substantia nigra, red nucleus, and caudate as previously highlighted in smaller studies,^9,14,63^ as well as several additional associations including three WM regions. Of these regions, the cerebellar dentate nucleus showed the strongest inverse correlation with survival. While the current study focused on total tau pathology burden, there may also be cell-type–specific contributions to survival with previous work comparing pathological features in long- and short-duration PSP patients indicating that disease progression and neurodegeneration may be more strongly influenced by oligodendroglial rather than neuronal tau pathology.^56^

Although previous studies suggest that co-pathologies do not influence PSP phenotype or progression,^12,13^ we observed that despite being infrequent overall, limbic TDP-43 co-pathology was present in the majority of PSP-SL patients. This observation may reflect shared molecular pathways, as proposed in corticobasal degeneration where TDP-43 co-pathology has been implicated in modifying clinical phenotype.^64^ In our cohort, PSP-SL cases were, on average, 5.7 years older at death than the mean cohort age, suggesting that TDP-43 co-pathology in these patients may alternatively be age-related. Data regarding sex-related differences in PSP have been inconsistent,^65^ with some studies indicating that male patients have shorter survival^21,48,54^ and that female patients have worse executive dysfunction and faster cognitive decline.^66^ While no sex-related differences in clinical progression were observed, the predominance of female patients in the PSP-CBS group may indicate differences in selective regional vulnerability, supported by nominally lower precentral gyrus tau pathology in female patients with PSP-CBS, implying that they may develop clinical symptoms despite less overall tau accumulation. In line with prior research, we did not find a significant association between *MAPT* haplotype or *APOE* genotype and survival,^28,67^ nor did we observe a significant association between the presence of the *SLC2A13* SNP rs2242367 and survival in our models.^31^ However, the *MAPT* H2 haplotype may be associated with either a less toxic misfolded tau strain or greater neuronal resilience to tau pathology, as suggested by the older age at symptom onset in *MAPT* H2 carriers despite higher tau pathology burden in several cortical and subcortical regions. This contrasts with a previous report showing reduced tau pathology among *MAPT* H2 carriers.^27^ Mildly increased overall neurofibrillary tangle severity has previously been reported in PSP patients carrying the *TRIM11* SNP rs564309; this effect was not region-specific, and no differences were observed for other tau inclusions, including tufted astrocytes and coiled bodies^30^. In the present study, although *TRIM11* (A) patients had nominally lower tau pathology burden in several limbic regions, no regional differences remained significant after correction for multiple comparisons.

Work combining resting-state fMRI connectomics with [¹⁸F]PI-2620 tau-PET and post-mortem regional tau pathology suggests that brain connectivity underlies stronger inter-regional correlations in tau burden measured both in vivo and post-mortem.^68^ Significant covariance in tau pathology observed between anatomically connected brain regions in the present study provides further evidence for inter-regional connectivity in the spreading of tau pathology. Strong correlations between tau pathology burden in distant brain regions implicate spread via long-range tracts such as the corticospinal pathway (white matter of superior/middle frontal gyri, centrum semiovale, internal capsule and cerebral peduncle) and frontostriatal projections (superior frontal cortex to caudate), as well as more localised cortico-cortical association fibres and the nigrostriatal pathway (substantia nigra to caudate). Strong correlations between tau pathology burden in the precentral gyrus GM and the red nucleus, as well as between the red nucleus and the dentate nucleus, support tau pathology spread along corticorubral and dentatorubral pathways, respectively. The association between middle temporal and fusiform regions may reflect AD co-pathology rather than a primary PSP-related process. These data could also be interpreted that functionally or structurally connected brain regions share intrinsic vulnerability factors as indicated by work combining AV-1451 tau PET imaging and resting state functional MRI showing that increased metabolic demand and a lack of trophic support are responsible for tau accumulation in PSP, rather than strong functional connectivity.^69^

The SuStaIn algorithm reconstructs progression patterns from cross-sectional data and has been applied to brain MRI, fluorodeoxyglucose-PET, and [18F]Florzolotau-PET data from PSP patients, each identifying two subtypes.^11,70,71^ MRI-SuStaIn identified a subcortical subtype with early brainstem, superior cerebellar peduncle and dentate nucleus atrophy and a cortical subtype with early frontal and insular atrophy alongside brainstem atrophy.^70^ Approximately 80% of patients with PSP-P, PSP-PGF and PSP-RS clinical phenotypes in this study were assigned to the MRI-SuStaIn subcortical subtype, and a similar proportion of patients with cortical phenotypes (PSP-CBS, PSP-SL, PSP-F) were assigned to the MRI-SuStaIn cortical subtype.^70^ FDG-PET-SuStaIn also identified a brainstem subtype with initial midbrain hypometabolism before progression to striatal and cortical structures, and a cortical subtype with early prefrontal hypometabolism preceding that in the caudate and midbrain structures.^71^ In contrast to MRI-SuStaIn, the majority of patients with a PSP-RS or PSP-P clinical phenotype were assigned to the cortical FDG-PET-SuStaIn subtype.^71^ [18F]Florzolotau-PET-SuStaIn revealed one subtype progressing from subcortical (red nucleus, subthalamic nucleus, raphe nucleus and globus pallidus) to cortical regions and another subtype with early simultaneous involvement of both subcortical and cortical brain regions.^11^ In this study patients were classified as having a PSP-RS or non-RS clinical phenotype, with 92% of the latter group comprised of PSP-P or PSP-PGF patients; 61% and 71% of patients with PSP-RS and non-RS clinical phenotypes were included in the early subcortical SuStaIn subtype.^11^

Our findings support a model of PSP tau progression wherein there are patients with early subcortical involvement followed by superior frontal gyrus WM and subsequent cortical GM involvement (subtype 1), whilst others have early simultaneous involvement of both frontal WM and subcortical regions (subtypes 2 and 3). The large proportion of cases with non-RS clinical phenotypes in the present study, along with granular quantitative data, enabled the separation of patients with early simultaneous involvement of subcortical and frontal WM into those with earlier posterior (subtype 2) or anterior (subtype 3) frontal involvement. The clinical phenotypes of patients in SuStaIn subtype 1, identified using quantitative tau pathology data, correspond closely with those of patients in the MRI-SuStaIn subcortical subtype, thus supporting a common progression pattern for most patients with PSP-RS and PSP-P. The agreement is weaker for the cortical subtype, which indicates that factors beyond regional tau pathology burden, including disrupted functional connectivity^72^ or intrinsic cellular vulnerability may contribute to clinical heterogeneity in these patients. However, differences in anatomical regions used for SuStaIn modelling across studies and the possible inclusion of patients with non-PSP pathology in imaging studies may also contribute to the observed discrepancy.

This work shares limitations common to brain bank post-mortem studies, including the potential for referral bias and non-standardised clinical assessments performed by different professionals. To limit this potential bias, only cases with detailed and complete information were included. Of note, autonomic function testing and polysomnography were not available to confirm autonomic symptoms and REM sleep behaviour disorder respectively, and we recorded all visual hallucinations irrespective of duration, or potential contribution of precipitating factors. A limitation regarding data-driven modelling of disease stage was the absence of early stage, incidental PSP patients with such cases being very uncommon in the QSBB cohort. Strengths include the large number of patients, the availability of genetic and detailed longitudinal clinical data throughout the entire disease course enabling standardised clinical phenotyping, and quantitative assessment of regional tau pathology using digital pathology methods, which reduce inter and intra-rater bias. In addition, by including only patients with pathologically confirmed PSP in the data-driven models, we were able to minimise the influence of AD co-pathology by excluding patients with AD pathology sufficient to affect the model output.

In summary, our results support a model in which PSP tau pathology spreads along anatomically connected networks, and via cell-autonomous mechanisms leading to the early emergence of PSP tau pathology in distant brain regions. This study also provides additional longitudinal clinical data on a large cohort of PSP cases, with novel anatomical reference standards for the expected regional distribution of tau pathology across MDS-defined PSP phenotypes, and characterises the relationship between tau pathology and survival, with translational implications for studies evaluating therapies targeting tau removal and *in vivo* tau imaging in PSP.^3,73^ Digital pathology methods together with advances in machine learning algorithms hold the potential for performing large-scale, unbiased, and cell-specific quantification of primary tau pathology and co-pathologies across multiple clinicopathological cohorts. Integration of such data with region- and cell-specific gene expression redouts and inclusion as quantitative phenotypic traits in genome-wide association studies will further improve our understanding of the molecular drivers of tau propagation patterns and phenotypic diversity in PSP.

## Acknowledgements

We thank the patients and their families, without whose generous donations this study would not have been possible. We also thank the QSBB administrative and technical staff for assistance in tissue preparation. We thank Jaime Anton Arnal for assistance with computing. A large language model (ChatGPT, OpenAI) was used to improve the clarity and conciseness of specific phrases and sentences in the manuscript. All suggestions were critically reviewed and revised by the authors to ensure the accuracy and integrity of the scientific content. Fig. 1 was created using BioRender.

## Funding

PWC is funded by the Reta Lila Weston Trust and the Edmond J. Safra Foundation. ALY and LPB are funded by the Wellcome Trust [227341/Z/23/Z]. EdPF is funded by a Senior Research Fellowship from Parkinson’s UK. For the purpose of open access, the author has applied a CC BY public copyright licence to any Author Accepted Manuscript version arising from this submission.

## Competing interests

The authors report no competing interests.

## Supplementary material

Supplementary material is available at *Brain* online.

## Notes

### Competing Interest Statement

The authors have declared no competing interest.

### Author Declarations

QSBB protocols and this study have been approved by the NHS Health Research Authority Ethics Committee London-Central (REC reference 23/LO/0044). All donors included in this study or their next of kin gave written informed consent.

## References

1. Roemer SF, Grinberg LT, Crary JF, et al. Rainwater Charitable Foundation criteria for the neuropathologic diagnosis of progressive supranuclear palsy. Acta Neuropathol. 2022;144:603–614.

2. Cullinane PW, Wrigley S, Bezerra Parmera J, et al. Pathology of neurodegenerative disease for the general neurologist. Pract Neurol. 2024;24:188–199.

3. Street D, Malpetti M, Rittman T, et al. Clinical progression of progressive supranuclear palsy: impact of trials bias and phenotype variants. Brain Commun. 2021;3:fcab206.

4. Street D, Jabbari E, Costantini A, et al. Progression of atypical parkinsonian syndromes: PROSPECT-M-UK study implications for clinical trials. Brain. 2023;146:3232–3242.

5. Kovacs GG, Lukic MJ, Irwin DJ, et al. Distribution patterns of tau pathology in progressive supranuclear palsy. Acta Neuropathol. 2020;140:99–119.

6. Tsuboi Y, Josephs KA, Boeve BF, et al. Increased tau burden in the cortices of progressive supranuclear palsy presenting with corticobasal syndrome. Mov Disord. 2005;20:982–988.

7. Ahmed Z, Josephs KA, Gonzalez J, DelleDonne A, Dickson DW. Clinical and neuropathologic features of progressive supranuclear palsy with severe pallido-nigro-luysial degeneration and axonal dystrophy. Brain. 2008;131:460–472.

8. Ling H, de Silva R, Massey LA, et al. Characteristics of progressive supranuclear palsy presenting with corticobasal syndrome: a cortical variant. Neuropathol Appl Neurobiol. 2014;40:149–163.

9. Williams DR, Holton JL, Strand C, et al. Pathological tau burden and distribution distinguishes progressive supranuclear palsy-parkinsonism from Richardson’s syndrome. Brain. 2007;130:1566–1576.

10. Sakae N, Josephs KA, Litvan I, et al. Neuropathologic basis of frontotemporal dementia in progressive supranuclear palsy. Mov Disord. 2019;34:1655–1662.

11. Hong J, Lu J, Liu F, et al. Uncovering distinct progression patterns of tau deposition in progressive supranuclear palsy using [18F]Florzolotau PET imaging and subtype/stage inference algorithm. EBioMedicine. 2023;97:104835.

12. Robinson JL, Yan N, Caswell C, et al. Primary Tau Pathology, Not Copathology, Correlates With Clinical Symptoms in PSP and CBD. J Neuropathol Exp Neurol. 2020;79:296–304.

13. Jecmenica Lukic M, Kurz C, Respondek G, et al. Copathology in Progressive Supranuclear Palsy: Does It Matter? Mov Disord. 2020;35:984–993.

14. Badihian N, Tosakulwong N, Weigand SD, et al. Relationships between regional burden of tau pathology and age at death and disease duration in PSP. Parkinsonism Relat Disord. 2024;127:107109.

15. Young AL, Marinescu RV, Oxtoby NP, et al. Uncovering the heterogeneity and temporal complexity of neurodegenerative diseases with Subtype and Stage Inference. Nat Commun. 2018;9:4273.

16. Wrigley S, Cullinane PW, Parmera JB, et al. Clinical Diagnosis of Progressive Supranuclear Palsy (PSP): A Clinicopathological Comparison of Patients with Confirmed PSP and Clinical Mimics. Mov Disord. Published online 12 June 2025. 10.1002/mds.30261

17. Höglinger GU, Respondek G, Stamelou M, et al. Clinical diagnosis of progressive supranuclear palsy: The movement disorder society criteria. Mov Disord. 2017;32:853–864.

18. Grimm MJ, Respondek G, Stamelou M, et al. How to Apply the Movement Disorder Society Criteria for Diagnosis of Progressive Supranuclear Palsy. Mov Disord. 2019;34:1228–1232.

19. Shoeibi A, Litvan I, Juncos JL, et al. Are the International Parkinson disease and Movement Disorder Society progressive supranuclear palsy (IPMDS-PSP) diagnostic criteria accurate enough to differentiate common PSP phenotypes? Parkinsonism Relat Disord. 2019;69:34–39.

20. Frank A, Peikert K, Linn J, Brandt MD, Hermann A. MDS criteria for the diagnosis of progressive supranuclear palsy overemphasize Richardson syndrome. Ann Clin Transl Neurol. 2020;7:1702–1707.

21. O’Sullivan SS, Massey LA, Williams DR, et al. Clinical outcomes of progressive supranuclear palsy and multiple system atrophy. Brain. 2008;131:1362–1372.

22. Bandres-Ciga S, Faghri F, Majounie E, et al. NeuroBooster Array: A Genome-Wide Genotyping Platform to Study Neurological Disorders Across Diverse Populations. Mov Disord. 2024;39:2039–2048.

23. Blauwendraat C, Faghri F, Pihlstrom L, et al. NeuroChip, an updated version of the NeuroX genotyping platform to rapidly screen for variants associated with neurological diseases. Neurobiol aging. 2017;57:247.e9.

24. Chang CC, Chow CC, Tellier LC, Vattikuti S, Purcell SM, Lee JJ. Second-generation PLINK: rising to the challenge of larger and richer datasets. GigaScience. 2015;4:7.

25. Taliun D, Harris DN, Kessler MD, et al. Sequencing of 53,831 diverse genomes from the NHLBI TOPMed Program. Nature. 2021;590:290–299.

26. Loh PR, Danecek P, Palamara PF, et al. Reference-based phasing using the Haplotype Reference Consortium panel. Nat Genet. 2016;48:1443–1448.

27. Heckman MG, Brennan RR, Labbé C, et al. Association of MAPT Subhaplotypes With Risk of Progressive Supranuclear Palsy and Severity of Tau Pathology. JAMA Neurol. 2019;76:710–717.

28. Baba Y, Putzke JD, Tsuboi Y, et al. Effect of MAPT and APOE on prognosis of progressive supranuclear palsy. Neurosci Lett. 2006;405:116–119.

29. Jabbari E, Woodside J, Tan MMX, et al. Variation at the TRIM11 locus modifies progressive supranuclear palsy phenotype. Ann Neurol. 2018;84:485–496.

30. Valentino RR, Koga S, Heckman MG, et al. Association of Tripartite Motif Containing 11 rs564309 With Tau Pathology in Progressive Supranuclear Palsy. Mov Disord. 2020;35:890–894.

31. Jabbari E, Koga S, Valentino RR, et al. Genetic determinants of survival in progressive supranuclear palsy: a genome-wide association study. Lancet Neurol. 2021;20:107–116.

32. Cicognola C, Salvadó G, Smith R, et al. APOE4 impact on soluble and insoluble tau pathology is mostly influenced by amyloid-β. Brain. 2025;148:2373–2383.

33. Wang H, Chang TS, Dombroski BA, et al. Whole-genome sequencing analysis reveals new susceptibility loci and structural variants associated with progressive supranuclear palsy. Mol Neurodegener. 2024;19:61.

34. Müller U, Höglinger G, Dickson DW. Multifactorial etiology of progressive supranuclear palsy (PSP): the genetic component. Acta Neuropathol. 2025;149:58.

35. McKeith IG, Galasko D, Kosaka K, et al. Consensus guidelines for the clinical and pathologic diagnosis of dementia with Lewy bodies (DLB): report of the consortium on DLB international workshop. Neurology. 1996;47:1113–1124.

36. Braak H, Braak E. Neuropathological stageing of Alzheimer-related changes. Acta Neuropathol. 1991;82:239–259.

37. Braak H, Del Tredici K, Rüb U, de Vos RAI, Jansen Steur ENH, Braak E. Staging of brain pathology related to sporadic Parkinson’s disease. Neurobiol Aging. 2003;24:197–211.

38. Thal DR, Rüb U, Orantes M, Braak H. Phases of Aβ-deposition in the human brain and its relevance for the development of AD. Neurology. 2002;58:1791–1800.

39. Hyman BT, Phelps CH, Beach TG, et al. National Institute on Aging–Alzheimer’s Association guidelines for the neuropathologic assessment of Alzheimer’s disease. Alzheimers Dement. 2012;8:1–13.

40. Briggs M, Allinson KSJ, Malpetti M, Spillantini MG, Rowe JB, Kaalund SS. Validation of the new pathology staging system for progressive supranuclear palsy. Acta Neuropathol. 2021;141:787–789.

41. Bankhead P, Loughrey MB, Fernández JA, et al. QuPath: Open source software for digital pathology image analysis. Sci Rep. 2017;7:16878.

42. Otsu N. A Threshold Selection Method from Gray-Level Histograms. *IEEE Transactions on Systems*, Man, and Cybernetics. 1979;9:62–66.

43. Montine TJ, Phelps CH, Beach TG, et al. National Institute on Aging-Alzheimer’s Association guidelines for the neuropathologic assessment of Alzheimer’s disease: a practical approach. Acta Neuropathol. 2012;123:1–11.

44. Respondek G, Stamelou M, Kurz C, et al. The phenotypic spectrum of progressive supranuclear palsy: a retrospective multicenter study of 100 definite cases. Mov Disord. 2014;29:1758–1766.

45. Williams DR, de Silva R, Paviour DC, et al. Characteristics of two distinct clinical phenotypes in pathologically proven progressive supranuclear palsy: Richardson’s syndrome and PSP-parkinsonism. Brain. 2005;128:1247–1258.

46. Litvan I, Mangone CA, McKee A, et al. Natural history of progressive supranuclear palsy (Steele-Richardson-Olszewski syndrome) and clinical predictors of survival: a clinicopathological study. J Neurol Neurosurg Psychiatry. 1996;60:615–620.

47. Glasmacher SA, Leigh PN, Saha RA. Predictors of survival in progressive supranuclear palsy and multiple system atrophy: a systematic review and meta-analysis. J Neurol Neurosurg Psychiatry. 2017;88:402–411.

48. Chiu WZ, Kaat LD, Seelaar H, et al. Survival in progressive supranuclear palsy and frontotemporal dementia. J Neurol Neurosurg Psychiatry. 2010;81:441–445.

49. Sánchez-Ruiz de Gordoa J, Zelaya V, Tellechea-Aramburo P, et al. Is the Phenotype Designation by PSP-MDS Criteria Stable Throughout the Disease Course and Consistent With Tau Distribution? Front Neurol. 2022;13:827338.

50. Nath U, Ben-Shlomo Y, Thomson RG, Lees AJ, Burn DJ. Clinical features and natural history of progressive supranuclear palsy: a clinical cohort study. Neurology. 2003;60:910–916.

51. dell’Aquila C, Zoccolella S, Cardinali V, et al. Predictors of survival in a series of clinically diagnosed progressive supranuclear palsy patients. Parkinsonism Relat Disord. 2013;19:980–985.

52. Papapetropoulos S, Gonzalez J, Mash DC. Natural history of progressive supranuclear palsy: a clinicopathologic study from a population of brain donors. Eur Neurol. 2005;54:1–9.

53. Oliveira MCB, Ling H, Lees AJ, Holton JL, De Pablo-Fernandez E, Warner TT. Association of autonomic symptoms with disease progression and survival in progressive supranuclear palsy. J Neurol Neurosurg Psychiatry. 2019;90:555–561.

54. Santacruz P, Uttl B, Litvan I, Grafman J. Progressive supranuclear palsy: a survey of the disease course. Neurology. 1998;50:1637–1647.

55. Xie T, Yuen CA, Kang W, Padmanaban M, Hain TC, Nichols J. Severity of Downgaze Palsy in the Context of Disease Duration Could Estimate Survival Duration in Patients With Progressive Supranuclear Palsy. Front Neurol. 2021;12:736784.

56. Lukic MJ, Respondek G, Kurz C, et al. Long-Duration Progressive Supranuclear Palsy: Clinical Course and Pathological Underpinnings. Ann Neurol. 2022;92:637–649.

57. Cosseddu M, Benussi A, Gazzina S, et al. Natural history and predictors of survival in progressive supranuclear palsy. J Neurol Sci. 2017;382:105–107.

58. Arena JE, Weigand SD, Whitwell JL, et al. Progressive supranuclear palsy: progression and survival. J Neurol. 2016;263:380–389.

59. Dickson DW, Ahmed Z, Algom AA, Tsuboi Y, Josephs KA. Neuropathology of variants of progressive supranuclear palsy. Curr Opin Neurol. 2010;23:394–400.

60. Koizumi R, Akagi A, Riku Y, et al. Correlation between clinical and neuropathological subtypes of progressive supranuclear palsy. Parkinsonism Relat Disord. 2024;127: 106076.

61. Williams DR, Holton JL, Strand K, Revesz T, Lees AJ. Pure akinesia with gait freezing: a third clinical phenotype of progressive supranuclear palsy. Mov Disord. 2007;22:2235–2241.

62. Whitwell JL, Tosakulwong N, Clark HM, et al. Diffusion tensor imaging analysis in three progressive supranuclear palsy variants. J Neurol. 2021;268:3409–3420.

63. Couto B, Martinez-Valbuena I, Lee S, et al. Protracted course progressive supranuclear palsy. Eur J Neurol. 2022;29:2220–2231.

64. Koga S, Kouri N, Walton RL, et al. Corticobasal degeneration with TDP-43 pathology presenting with progressive supranuclear palsy syndrome: A distinct clinicopathologic subtype. Acta Neuropathol. 2018;136:389–404.

65. Digma LA, Litvan I, del Ser T, Bayram E. Sex differences for cognitive decline in progressive supranuclear palsy. Parkinsonism Relat Disord. 2023;112:105454.

66. Mahale RR, Krishnan S, Divya KP, Jisha VT, Kishore A. Gender differences in progressive supranuclear palsy. Acta Neurol Belg. 2022;122:357–362.

67. Litvan I, Baker M, Hutton M. Tau genotype: no effect on onset, symptom severity, or survival in progressive supranuclear palsy. Neurology. 2001;57:138–140.

68. Franzmeier N, Brendel M, Beyer L, et al. Tau deposition patterns are associated with functional connectivity in primary tauopathies. Nat Commun. 2022;13:1362.

69. Cope TE, Rittman T, Borchert RJ, et al. Tau burden and the functional connectome in Alzheimer’s disease and progressive supranuclear palsy. Brain. 2018;141:550–567.

70. Scotton WJ, Shand C, Todd E, et al. Uncovering spatiotemporal patterns of atrophy in progressive supranuclear palsy using unsupervised machine learning. Brain Commun. 2023;5:fcad048.

71. Wang H, Wang B, Liao Y, et al. Identification of metabolic progression and subtypes in progressive supranuclear palsy by PET molecular imaging. Eur J Nucl Med Mol Imaging. 2024;52:823–835.

72. Sintini I, Ali F, Stephens Y, et al. Functional connectivity abnormalities in clinical variants of progressive supranuclear palsy. NeuroImage: Clinical. 2025;45:103727.

73. Marotta C, Sinclair B, O’Brien TJ, Vivash L. Biomarkers of disease progression in progressive supranuclear palsy for use in clinical trials. Brain Commun. 2025;7:fcaf022.

