## Supplementary Data for "Data-driven modelling of tau pathology reveals distinct progressive supranuclear palsy subtypes"

**Supplementary Table 1 Abbreviations and names of brain regions of interest**

| Region of interest | | % missing | Region of interest | | % missing |
|---|---|---|---|---|---|
| SFG-GM[a] | Superior frontal gyrus GM | 0.4 | CA1 | Cornu ammonis subregion 1 | 0.8 |
| SFG-WM[a] | Superior frontal gyrus WM | 0.4 | CA2 | Cornu ammonis subregion 2 | 0.8 |
| MFG-GM | Middle frontal gyrus GM | 0.4 | CA3 | Cornu ammonis subregion 3 | 1.3 |
| MFG-WM | Middle frontal gyrus WM | 0.4 | CA4 | Cornu ammonis subregion 4 | 0.4 |
| PCG-GM[a] | Precentral gyrus GM | 1.3 | DENT | Dentate gyrus | 0.4 |
| PCG-WM[a] | Precentral gyrus WM | 1.3 | GLOB_PALL[a] | Globus pallidus | 0.4 |
| SPL-GM[a] | Superior parietal lobule GM | 0.4 | CAU[a] | Caudate | 0.8 |
| SPL-WM[a] | Superior parietal lobule WM | 0.4 | INT-CAP[a] | Internal capsule | 0.4 |
| IPL-GM | Inferior parietal lobule GM | 0.4 | PUT | Putamen | 0.4 |
| IPL-WM | Inferior parietal lobule WM | 0.4 | DEEP-WM | Centrum Semiovale WM | 5.9 |
| STG-GM | Superior temporal gyrus GM | 0.4 | LAT-CP | Lateral cerebral peduncle | 6.8 |
| STG-WM | Superior temporal gyrus WM | 0.4 | CENT-CP | Central cerebral peduncle | 6.8 |
| MTG-GM[a] | Middle temporal gyrus GM | 0.4 | MED-CP | Medial cerebral peduncle | 8.5 |
| MTG-WM[a] | Middle temporal gyrus WM | 0.4 | SUB_NIGRA[a] | Substantia nigra | 4.2 |
| FG-GM | Fusiform gyrus GM | 0.8 | TEGMENT[a] | Midbrain tegmentum | 5.1 |
| FG-WM | Fusiform gyrus WM | 3.0 | RED_NUC[a] | Red nucleus | 5.9 |
| ENTC | Entorhinal cortex | 0.4 | SUP_COLL | Superior colliculus | 8.5 |
| TRANSENTC | Transentorhinal cortex | 0.4 | PERIV-GM | Periventricular GM | 10.2 |
| PARASUB | Parasubiculum | 1.3 | CBM-DN | Cerebellar dentate nucleus | 0.4 |
| SUB | Subiculum | 0.8 | CBM-WM[a] | Cerebellar WM | 0.4 |
| PHG-WM | Parahippocampal gyrus WM | 0.4 | | | |

[a]Regions of interest used in SuStaIn analyses.

**Supplementary Table 2 Brain region composites and constituent regions of interest**

| Brain region composites and constituent regions of interest | |
|---|---|
| Cortical GM | SFG-GM, MFG-GM, PCG-GM, SPL-GM, IPL-GM, STG-GM, MTG-GM |
| Cortical WM | SFG-WM, MFG-WM, PCG-WM, SPL-WM, IPL-WM, STG-WM, MTG-WM |
| Limbic GM | FG-GM, ENTC, TRANSENTC, PARASUB, SUB, CA1, CA2, CA3, CA4, DENT |
| Limbic WM | FG-WM, PHG-WM |
| Basal ganglia | GLOB_PALL, CAU, PUT |
| Deep WM | INT-CAP, DEEP-WM |
| Midbrain GM | TEGMENT, RED_NUC |
| Cerebral peduncle | LAT-CP, CENT-CP, MED-CP |
| Dentate nucleus | CBM-DN |
| Cerebellar WM | CBM-WM |

**Supplementary Table 3 Region-wise associations between tau pathology and survival**

| Region | β | *P*-value | *P* adj. | Region | β | *P*-value | *P* adj. |
|---|---|---|---|---|---|---|---|
| SFG-GM | -0.01171 | 0.092769 | 0.185539 | CA1 | 0.002766 | 0.206177 | 0.318586 |
| SFG-WM | -0.01101 | 0.310043 | 0.37581 | CA2 | 0.004821 | 0.087032 | 0.183225 |
| MFG-GM | -0.01715 | 0.035094 | 0.093585 | CA3 | 0.009424 | 0.039444 | 0.09861 |
| MFG-WM | -0.01824 | 0.215046 | 0.318586 | CA4 | 0.009247 | 0.342997 | 0.393224 |
| PCG-GM | -0.00414 | 0.286932 | 0.358665 | DENT | 0.00448 | 0.357787 | 0.397541 |
| PCG-WM | -0.02277 | 0.008258 | 0.030031 | GLOB_PALL | -0.02095 | 6.17E-05 | 0.001233 |
| SPL-GM | -0.00734 | 0.223652 | 0.318815 | CAU | -0.02076 | 0.002483 | 0.014189 |
| SPL-WM | -0.02936 | 0.011761 | 0.039204 | INT-CAP | -0.0114 | 0.034656 | 0.093585 |
| IPL-GM | 0.000512 | 0.950055 | 0.950055 | PUT | -0.0178 | 0.001838 | 0.012254 |
| IPL-WM | -0.03635 | 0.107513 | 0.200896 | DEEP-WM | -0.01957 | 0.007356 | 0.030031 |
| STG-GM | 0.004874 | 0.412303 | 0.441946 | LAT-CP | -0.01391 | 0.279195 | 0.358665 |
| STG-WM | 0.020751 | 0.213612 | 0.318586 | CENT-CP | -0.0092 | 0.419849 | 0.441946 |
| MTG-GM | 0.011128 | 0.043159 | 0.101549 | MED-CP | 0.008533 | 0.23538 | 0.318815 |
| MTG-WM | 0.037602 | 0.178489 | 0.310416 | SUB_NIGRA | -0.01914 | 0.005008 | 0.025038 |
| FG-GM | 0.006965 | 0.034184 | 0.093585 | TEGMENT | -0.01433 | 0.001291 | 0.01175 |
| FG-WM | 0.017297 | 0.239111 | 0.318815 | RED_NUC | -0.01434 | 0.000201 | 0.002675 |
| ENTC | 0.003244 | 0.110493 | 0.200896 | SUP_COLL | -0.0102 | 0.00816 | 0.030031 |
| TRANSENTC | 0.002677 | 0.187507 | 0.312511 | PERIV-GM | -0.01738 | 0.001469 | 0.01175 |
| PARASUB | 0.006787 | 0.056159 | 0.124797 | CBM-DN | -0.06197 | 5.54E-05 | 0.001233 |
| SUB | 0.00246 | 0.126187 | 0.26499 | CBM-WM | -0.0081 | 0.496518 | 0.509249 |
| PHG-WM | 0.005736 | 0.344071 | 0.393224 | | | | |

This table presents the results of Gamma generalized linear models (GLMs) assessing the association between regional tau pathology and survival. Models were conducted separately for each region. All models were adjusted for age at onset, sex, clinical phenotype at 3 years, and NIA-ABC score. For each region, beta coefficient (β), raw p-value, and Benjamini–Hochberg (BH) adjusted *P*-value are shown.

**Supplementary Table 4 Association between regional tau pathology composites and ages at onset and death**

| Tau pathology regional | Age at death ρ | *P*-value | Age at onset ρ | *P*-value |
|---|---|---|---|---|
| Cortical-GM | -0.051 | .468 | 0.026 | .710 |
| Limbic-GM | 0.235*** | <.001 | 0.257*** | <.001 |
| Basal ganglia-GM | -0.088 | .205 | 0.020 | .769 |
| Midbrain-GM | -0.273*** | <.001 | -0.186** | .007 |
| Dentate nucleus-GM | -0.031 | .654 | 0.097 | .164 |
| Cortical-WM | -.04 | .568 | 0.045 | .521 |
| Limbic-WM | 0.217** | .002 | 0.232*** | <.001 |
| Centrum semiovale-WM | -0.248*** | <.001 | -0.176* | .011 |
| Cerebral peduncle-WM | -0.056 | .419 | -0.051 | .458 |
| Cerebellum-WM | -0.143* | .039 | -0.102 | .141 |

Spearman correlations between age-related variables and regional tau pathology composites, after covariate adjustment. All variables were rank-transformed and adjusted for sex, NIA-ABC score, and clinical phenotype using linear models. Spearman correlation coefficients were computed between the residuals. Asterisks denote significance levels: $P < 0.05$ (*), $P < 0.01$ (**), $P < 0.001$ (***).

**Supplementary Table 5 Clinical, genetic, and neuropathological characteristics across SuStaIn subtypes**

| | Subtype 1 | Subtype 2 | Subtype 3 | *P*-value |
|---|---|---|---|---|
| **Female sex (no., %)** | 57 (83.8%) | 6 (8.8%) | 5 (7.4%) | 0.536 |
| **Age at onset (y)** | 67.62 ± 7.42 | 70.93 ± 7.72 | 67.74 ± 5.99 | 0.180 |
| **Survival (y)** | 7.47 ± 2.80 | 6.90 ± 2.26 | 7.15 ± 2.47 | 0.648 |
| **Age at death (y)** | 75.08 ± 7.19 | 77.84 ± 7.96 | 74.90 ± 6.08 | 0.279 |
| **NIA-ABC Score (no., %)** | | | | 0.228 |
| 0 | 50 (79.4%) | 8 (12.7%) | 5 (7.9%) | |
| 1 | 89 (80.2%) | 8 (7.2%) | 14 (12.6%) | |
| 2 | 10 (76.9%) | 3 (23.1%) | 0 (0) | |
| **Braak Stage (no., %)** | | | | 0.633 |
| 0 | 131 (78.9%) | 17 (10.2%) | 18 (10.8%) | |
| 1 | 1 (50.0%) | 1 (50.0%) | 0 (0) | |
| 2 | 2 (66.7%) | 1 (33.3%) | 0 (0) | |
| 3 | 3 (75.0%) | 0 (0) | 1 (25.0%) | |
| 4 | 6 (100.0%) | 0 (0) | 0 (0) | |
| 5 | 4 (100.0%) | 0 (0) | 0 (0) | |
| 6 | 2 (100.0%) | 0 (0) | 0 (0) | |
| **Amygdala-predominant LBD (no., %)** | | | | 0.542 |
| No | 140 (78.7%) | 19 (10.7%) | 19 (10.7%) | |
| Yes | 9 (100.0%) | | | |
| **TDP-43 proteinopathy (no., %)** | | | | 0.471 |
| No | 131 (80.4%) | 16 (9.8%) | 16 (9.8%) | |
| Yes | 15 (71.4%) | 3 (14.3%) | 3 (14.3%) | |
| ***APOE* ε4 status (no., %)** | | | | 0.359 |
| No | 97 (79.5%) | 15 (12.3%) | 10 (8.2%) | |
| Yes | 24 (77.4%) | 2 (6.5%) | 5 (16.1%) | |
| ***MAPT* H2 haplotype (no., %)** | | | | 0.505 |
| No | 110 (78.6%) | 15 (10.7%) | 15 (10.7%) | |
| Yes | 11 (84.6%) | 2 (15.4%) | | |
| ***TRIM11* rs564309 (A) (no., %)** | | | | |
| No | 103 (80.5%) | 15 (11.7%) | 10 (7.8%) | 0.192 |
| Yes | 18 (72.0%) | 2 (8.0%) | 5 (20.0%) | |
| ***SLC2A13* rs2242367 (A) (no., %)** | | | | |
| No | 72 (80.0%) | 9 (10.0%) | 9 (10.0%) | 0.872 |
| Yes | 49 (77.8%) | 8 (12.7%) | 6 (9.5%) | |

Continuous variables are summarised as mean ± standard deviation and compared using one-way ANOVA. Categorical variables are displayed as number of patients (percentage) for each level and comparisons were performed using chi-square or Fisher's exact tests where appropriate.

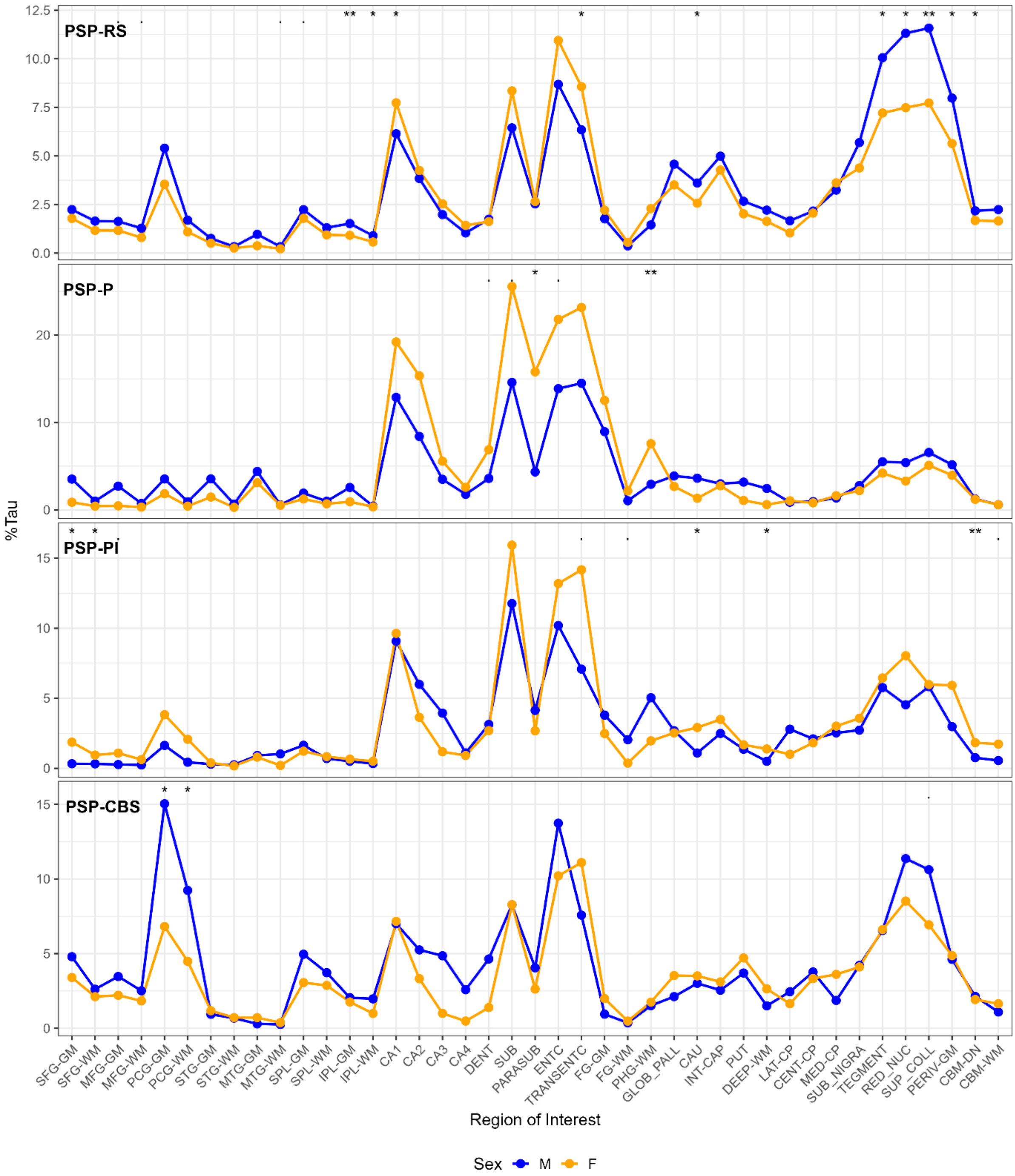


**Supplementary Figure 1 Sex differences in regional tau pathology burden by phenotype.** Line plots show mean tau pathology (%Tau) by region for males (blue) and females (orange). Each panel represents one subgroup. Asterisks indicate unadjusted pairwise Wilcoxon rank-sum test significance: ($P < 0.05$: *, $P < 0.01$: **). No comparisons survived Benjamini–Hochberg multiple testing correction.

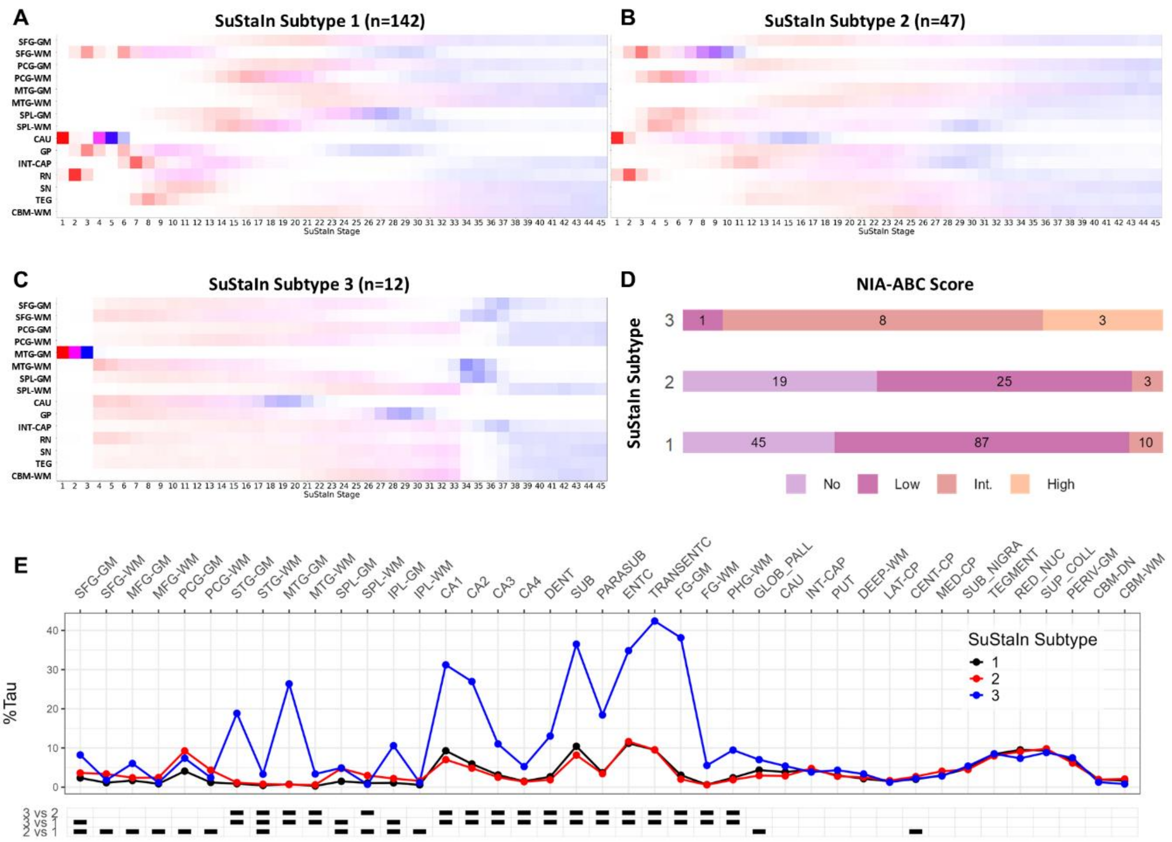


**Supplementary Figure 2 Preliminary Subtype and Stage Inference modelling of all PSP patients. (A-C)** Positional variance diagrams show the progression patterns of regional tau pathology inferred by the SuStaIn model for each of the three data-driven subtypes. Each row corresponds to a specific brain region, and each column to a SuStaIn stage; colour intensity reflects the probability that a given brain region is affected at a specific stage, with red indicating higher probability, blue lower probability, and white indicating low or no probability. **(D)** The bar chart shows the distribution NIA-ABC scores across SuStaIn subtypes. **(E)** The line plot shows the mean percentage of tau pathology (%Tau) across brain regions for each SuStaIn subtype. The tile plot below displays results from pairwise Wilcoxon rank-sum tests comparing subtypes at each region. Black squares indicate statistically significant differences after Bonferroni correction.

**Clinical features and disease progression milestone[1,2,3,4]**

**Alien limb:** The date of onset/first documentation of symptoms/signs of alien limb phenomenon.
**Apathy:** The date of onset/first documentation of reduced level of interest, initiative, and spontaneous activity; clearly apparent to informant or patient.
**Apraxia**: The date of onset/first documentation of orobuccal or limb apraxia.
**Axial rigidity:** The date of onset/first documentation of symptoms of axial rigidity or axial rigidity on examination.
**Behavioural change:** The date of onset/first documentation of impulsivity, disinhibition, or perseveration, e.g. socially inappropriate behaviours, overstuffing the mouth when eating, motor recklessness, applause sign, palilalia, echolalia.
**Bradykinesia/hypokinesia:** The date of onset/first documentation of symptoms of bradykinesia or bradykinesia/hypokinesia on examination.
**Bradyphrenia:** The date of onset/first documentation of slowed thinking; clearly apparent to informant or patient.
**Blepharospasm or Apraxia of Eyelid Opening**: The date of onset/first documentation of blepharospasm/AEO during either on physical examination or based on symptoms described by the patient or relative/carer.
**Cerebellar signs:** The date of first documentation of cerebellar signs on examination.
**Cortical sensory signs:** The date of onset/first documentation of cortical sensory deficits e.g. agraphesthesia, astereognosis.
**Dementia:** The date of onset/first documentation of cognitive impairment which affects the patient's ability to perform tasks of daily living.
**Diplopia/blurred vision:** The date of onset/first documentation of blurred vision or diplopia not explained by another cause.
**Dysexecutive syndrome:** The date of onset/first documentation of symptoms or signs of executive dysfunction including reverse digit span, Trails B or Stroop test, Luria sequence.
**Dysarthria:** The date of dysarthria documented by clinician or referral to Speech and Language therapy because of Speech impairment.
**Dysphagia:** The date of dysphagia sufficient to require referral to SLT for swallowing assessment, diet modification or of aspiration pneumonia.
**Dystonia:** The date of onset/first documentation of symptoms of dystonia or dystonia observed on examination, including craniofacial, cervical or Limb dystonia.

**Falls:** The date when the patient has reached the milestone of ≥2 falls/year or documentation of "frequent" or "regular" falls.

**Frontalis overactivity:** The date of first documentation of frontalis overactivity by clinician.

**Gait freezing:** The date of onset/first documentation of symptoms of gait freezing (upon gait initiation, turning etc…) or gait freezing observed on examination.

**Limb rigidity:** The date of onset/first documentation of symptoms of limb rigidity or limb rigidity on examination.

**Mild cognitive impairment:** The date of onset/first documentation of cognitive symptoms that does not interfere with daily activities.

**Myoclonus:** The date of onset/first documentation of myoclonus or myoclonus observed on examination.

**Orthostatic hypotension:** date of onset of symptomatic orthostatic hypotension or asymptomatic postural drop in blood pressure ≥20/10mm/Hg.

**Primary progressive aphasia:** The date of onset/first documentation of progressive language problems including loss of grammar and/or telegraphic speech or writing progressive aphasia or effortful, halting speech with inconsistent speech sound errors and distortions or slow syllabically segmented prosodic speech pattern with spared single-word comprehension, object knowledge, and word retrieval during sentence repetition.

**Pyramidal signs:** The date of first documentation of pyramidal signs on examination.

**Reduced verbal fluency:** The date of onset/first documentation of problems with verbal fluency either reported by the patient/carer or documented on examination.

**REM sleep behaviour disorder:** The date of onset/first documentation of body movement, emotional expression, or audible verbalisation of dream content during sleep time.

**Severe dysarthria:** The date when the patient's speech becomes unintelligible or requirement of communication aids.

**Severe dysphagia (PEG):** The date the patient was recommended to have a PEG/had PEG performed if former not available. In the event PEG tube placement was declined by patient, record the date it was recommended.

**Tremor:** The date of onset/first documentation of any rest, action, postural tremor either reported by the patient/carer or observed on examination.

**Urinary catheter:** The date the patient began to use a urinary catheter regularly.

**Urinary dysfunction**: The date of onset of lower urinary tract symptoms, including urinary urgency, frequency, or nocturia, incontinence.

**Vertical supranuclear gaze palsy (vSNP)**: The date of onset/first documentation of the first evidence of supranuclear gaze palsy (e.g. documented vertical supranuclear gaze palsy [O1] or slow velocity of vertical saccades [O2])

**Visual Hallucination:** The date of onset/first documentation of visual hallucination.

**References:**

1. Grimm M-J, Respondek G, Stamelou M, et al. How to Apply the Movement Disorder Society Criteria for Diagnosis of Progressive Supranuclear Palsy. *Mov Disord* 2019; 34: 1228–32.

2. Höglinger GU, Respondek G, Stamelou M, et al. Clinical diagnosis of progressive supranuclear palsy: The movement disorder society criteria. *Mov Disord* 2017; 32: 853–64.

3. Jecmenica Lukic M, Kurz C, Respondek G, et al. Copathology in Progressive Supranuclear Palsy: Does It Matter? *Mov Disord* 2020; 35: 984–93.

4. O'Sullivan SS, Massey LA, Williams DR, et al. Clinical outcomes of progressive supranuclear palsy and multiple system atrophy. *Brain* 2008; 131: 1362–72.
